# Projections and fractional dynamics of COVID-19 with optimal control analysis

**DOI:** 10.1101/2020.11.17.20233031

**Authors:** Khondoker Nazmoon Nabi, Pushpendra Kumar, Vedat Suat Erturk

## Abstract

When the entire world is eagerly waiting for a safe, effective and widely available COVID-19 vaccine, unprecedented spikes of new cases are evident in numerous countries. To gain a deeper understanding about the future dynamics of COVID-19, a compartmental mathematical model has been proposed in this paper incorporating all possible non-pharmaceutical intervention policies. Model parameters have been calibrated using sophisticated trust-region-reflective algorithm and short-term projection results have been illustrated for Argentina, Bangladesh, Brazil, Colombia and India. Control reproduction numbers (*ℛ*_*c*_) have been calculated in order to get insights about the current epidemic scenario in the above-mentioned countries. Forecasting results depict that the aforesaid countries are having downward trends in daily COVID-19 cases. However, it is highly recommended to use efficacious face coverings and maintain strict physical distancing, as the pandemic is not over in any country. Global sensitivity analysis enlightens the fact that efficacy of face coverings is the most significant parameter, which could significantly control the transmission dynamics of the novel coronavirus compared to other non-pharmaceutical measures. In addition, reduction in effective contact rate with isolated patients is also essential in bringing down the epidemic threshold (*ℛ*_*c*_) below unity. All necessary graphical simulations have been performed with the help of Caputo-Fabrizio fractional derivatives. In addition, optimal control problem for fractional system has been designed and the existence of unique solution has also been showed by using Picard-Lindelof technique. Finally, the unconditionally stability of the given fractional numerical technique has been proved.

## 1. Introduction

To date, as there is no world-wide accepted vaccine that can provide full immunity to the human body against COVID-19, non-pharmaceutical intervention strategies are the realistic and effective solutions to control second wave of the pandemic. However, around 40 different coronavirus vaccines are experiencing critical clinical trials and nine already in the final stage of testing on thousands of people. Importantly, a leading vaccine candidate developed by the University of Oxford is already in an advanced stage of testing and it has been found in trials that it can trigger an immune response [1]. Generally, an effective vaccine would take years, if not decades, to develop. As research in this field is happening at a breakneck speed, scientists believe that an effective vaccine is likely to become widely available by mid-2021. Lack of transparency could be a vital issue in upcoming days and a false sense of security could evolve among general people if the vaccine doesn’t work effectively.

Mathematical models can always provide considerable insights of the transmission dynamics and complexities of any infectious diseases, which eventually help government officials design overall epidemic planning. Importantly, mathematical analysis always plays a notable role in making vital public health decisions, resource allocation and implementation of social distancing measures and other non-pharmaceutical interventions. From the beginning of the COVID-19 outbreak, mathematicians and researchers are working relentlessly and have already done tremendous contributions in limiting the spread of the coronavirus in different parts of the world [2, 3, 4, 5, 6]. In an early contribution, Ferguson *et al*. [2] showed the impact of different non-pharmaceutical intervention strategies on COVID-19 mortality by developing an agent-based model. In another study, Ngonghala *et al.* showed that effective and comprehensive usage of face coverings can significantly limit the spread of the virus and reduce the COVID-induced mortality in different states of the USA in general in the absence of community lockdown measures and stringent social distancing practice. On the other hand, Nabi [5] projected the future dynamics of COVID-19 for various COVID-19 hotspots by proposing a compartmental mathematical model and concluded that early relaxation of lockdown measures and social distancing could bring a second wave in no time. As a matter of reality, inhabitants in several countries compelled to violate containment measures due to prolonged lockdown measures and severe economic recession [4]. Netherlands having one of the best health care systems in the world, is grappling with continuous spikes in daily cases due to aversion to masks.

In this study, in the absence of a safe, effective and world-wide approved vaccine, a new compartmental mathematical COVID-19 model has been designed incorporating all possible non-pharmaceutical intervention strategies such as wearing face coverings, social distancing, home or self-quarantine and self or institutional isolation. In addition, the impacts of different interventions scenarios have been analysed rigorously. The aim of this work is to project the future dynamics of COVID-19 outbreak in five countries namely Argentina, Bangladesh, Brazil, Colombia and India which are worst-hit countries in the world. Estimation of parameters has been performed by using real-time data, followed by a projection of the evolution of the disease. Global sensitivity analysis is applied to determine the influential mechanisms in the model that drive the transmission dynamics of the disease. For fractional simulations, we used the well known non-integer order derivative called Caputo-Fabrizio (CF) fractional derivative. Since last few decades, there are so many epidemic models have been solved by non-integer order derivatives. Recently some applications of non-integer order derivatives in mathematical epidemiology can be seen from [7, 8, 9, 10, 11]. There are so many research papers have been come to study the outbreaks of coronavirus, in which some are [12, 13, 14, 15, 16, 17, 18, 19, 20, 21]. We performed the optimal control problem in CF derivative sense and provided the existence of unique solution by well-known technique named as Picard-Lindelof technique. We also proved the unconditionally stability of the given fractional numerical technique. We used the numerical data of all five given countries and perform the all necessary graphical simulations.

The entire chapter is organized as follows. Materials and methods are presented in Section 2. Section 3 is solely devoted to asymptotic stability of the proposed model. In section 4, estimation of model parameters and projection results have been discussed using daily COVID-19 data of Argentina, Bangladesh, Brazil, Colombia and India. In section 5, to quantify the impact of different model mechanisms, LHS-PRCC global sensitivity scheme has been performed. In section 6, numerical and graphical simulations have been illustrated using Caputo-Fabrizio fractional derivatives. Later, optimal control problem has been designed in fractional sense in section 7. The chapter ends with some insightful findings and strategies, which could significantly control the transmission dynamics of COVID-19.

## 2. Materials and methods

### 2.1. Mode formulation

A compartmental mathematical has been designed to describe the transmission dynamics of the COVID-19 incorporating all possible real-life interactions. Considering different infection status, the entire human population (denoted by *N* (*t*) at time *t*) has been stratified into nine mutually-exclusive compartments of susceptible individuals (*S*(*t*)), early-exposed individuals (*E*_1_(*t*)), pre-symptomatic individuals (*E*_2_(*t*)), symptomatically-infectious (*I*(*t*)), asymptomatically-infectious or infectious individuals with mild-symptoms (*A*(*t*)), quarantined infectious (*Q*(*t*)), hospitalised or isolated individuals (*L*(*t*)), recovered individuals (*R*(*t*)), disease-induced death cases (*D*(*t*)). Hence,

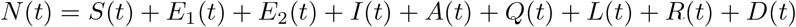

The following assumptions have been considered to formulate the above-mentioned model.

- Vital dynamic (birth and natural deaths) has been ignored as the main objective of this study is to observe the short-term dynamics of COVID-19 pandemic.
- Recovered individuals are immune to the disease which means that they cannot get reinfected. However, in a recent study of Tillett *et al.* [22], genomic evidence for reinfection with SARS-CoV-2 has been found. Nevertheless, it is important to note this singular finding does not provide generalisability of this phenomenon and hence it is not considered in our study.

The flow diagram of the proposed model is illustrated in Figure 1, where susceptible individuals can become infected by an effective contact with individuals in the pre-symptomatic (*E*_2_(*t*)), symptomatically-infectious (*I*(*t*)), asymptomatically-infectious (*A*(*t*)), quarantined-infectious (*Q*(*t*)) and isolated-infectious (*L*(*t*)). Effective contact rates are 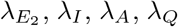, and *λ*_*L*_ respectively and the expressions are defined in (2). Importantly, the compartment *E*_1_(*t*) consists of early-infected individuals who are still not infectious, whereas the individuals in pre-symptomatic cohort *E*_2_(*t*) have the capability of transmitting coronavirus before the end of the disease incubation period. A proportion of individuals in newly-exposed compartment (*E*_1_(*t*)) progress to pre-symptomatic class (*E*_2_(*t*)) at a rate *κ*_1_. After the completion of disease mean incubation period, at a rate *ρκ*_2_, a fraction of individuals who have clear clinical symptoms of COVID-19 progress to *I*(*t*) compartment. Individuals in *E*_2_(*t*) class who do not have any clear symptoms progress to *A*(*t*) class at a rate (1 *− ρ*)*κ*_2_. Pre-symptomatic individuals are assumed to be self-quarantined at a rate *q*. With the help of diagnostic or surveillance testing approaches, symptomatically-infectious individuals and asymptomatically-infectious individuals are brought under institutional or home isolation at rates *τ*_*A*_ and *τ*_*I*_ respectively. Moreover, the parameter *γ*_*I*_ (*γ*_*A*_)(*γ*_*Q*_)(*γ*_*L*_) represents the recovery rate for individuals in the *I*(*A*)(*Q*)(*L*) class. Finally, the disease-induced mortality rate for individuals in the *I*(*Q*)(*L*) compartment is defined by the parameter *δ*_*I*_ (*δ*_*Q*_)(*δ*_*L*_). Considering all the above-mentioned interactions, the transmission dynamics of COVID-19 can be described by the following system of nonlinear ordinary differential equations.

**Figure 1:**
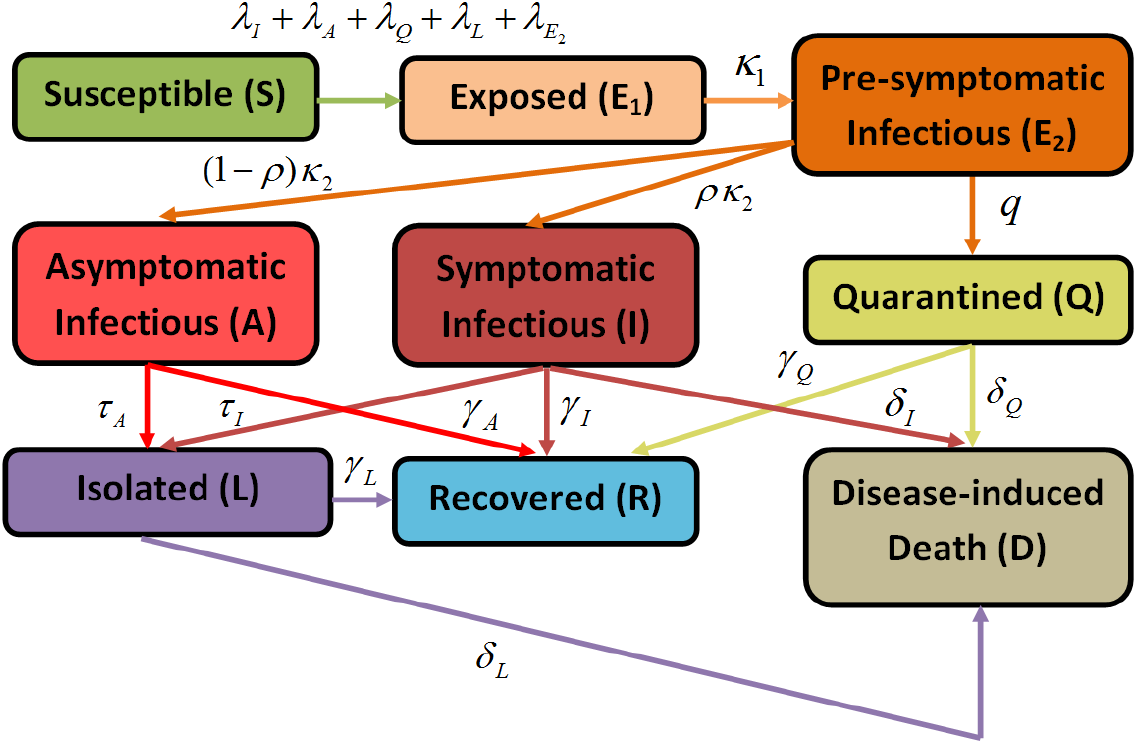
Flow diagram of the COVID-19 transmission dynamics.

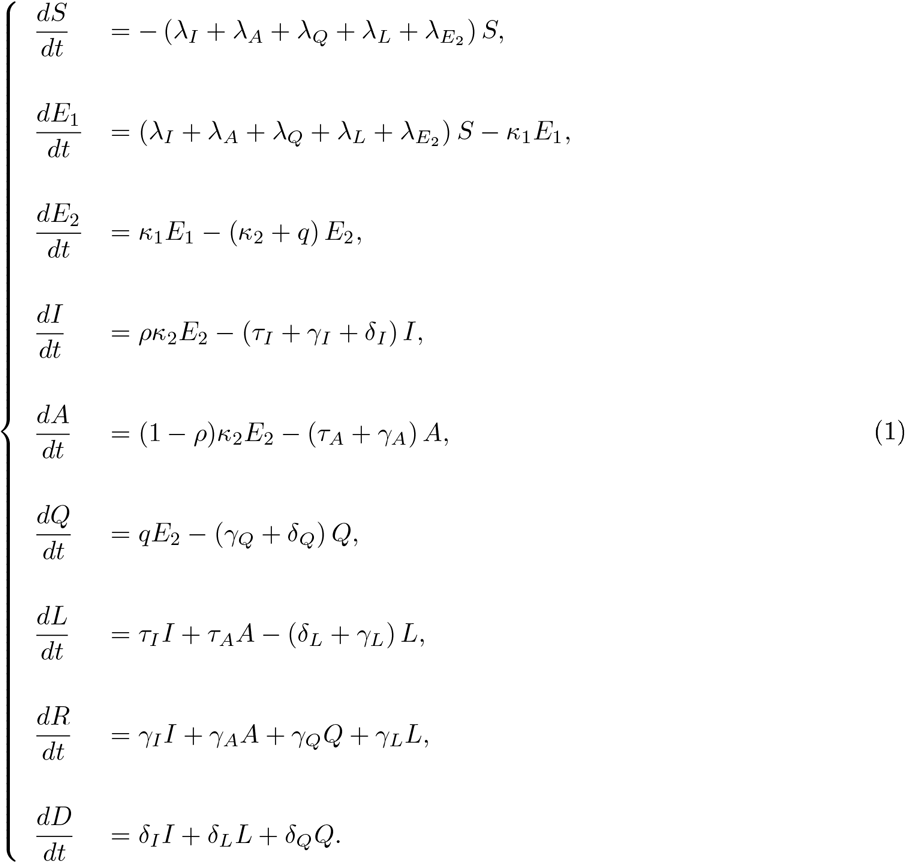

where the forces of infection are defined below

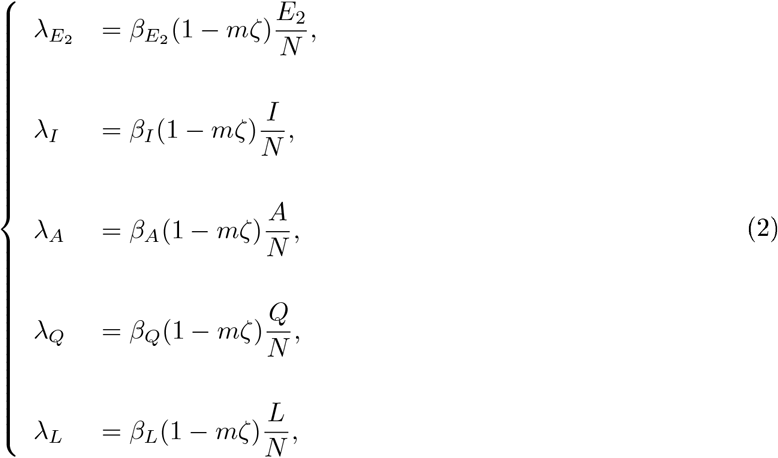

The parameters are described in Table 1.

**Table 1:**
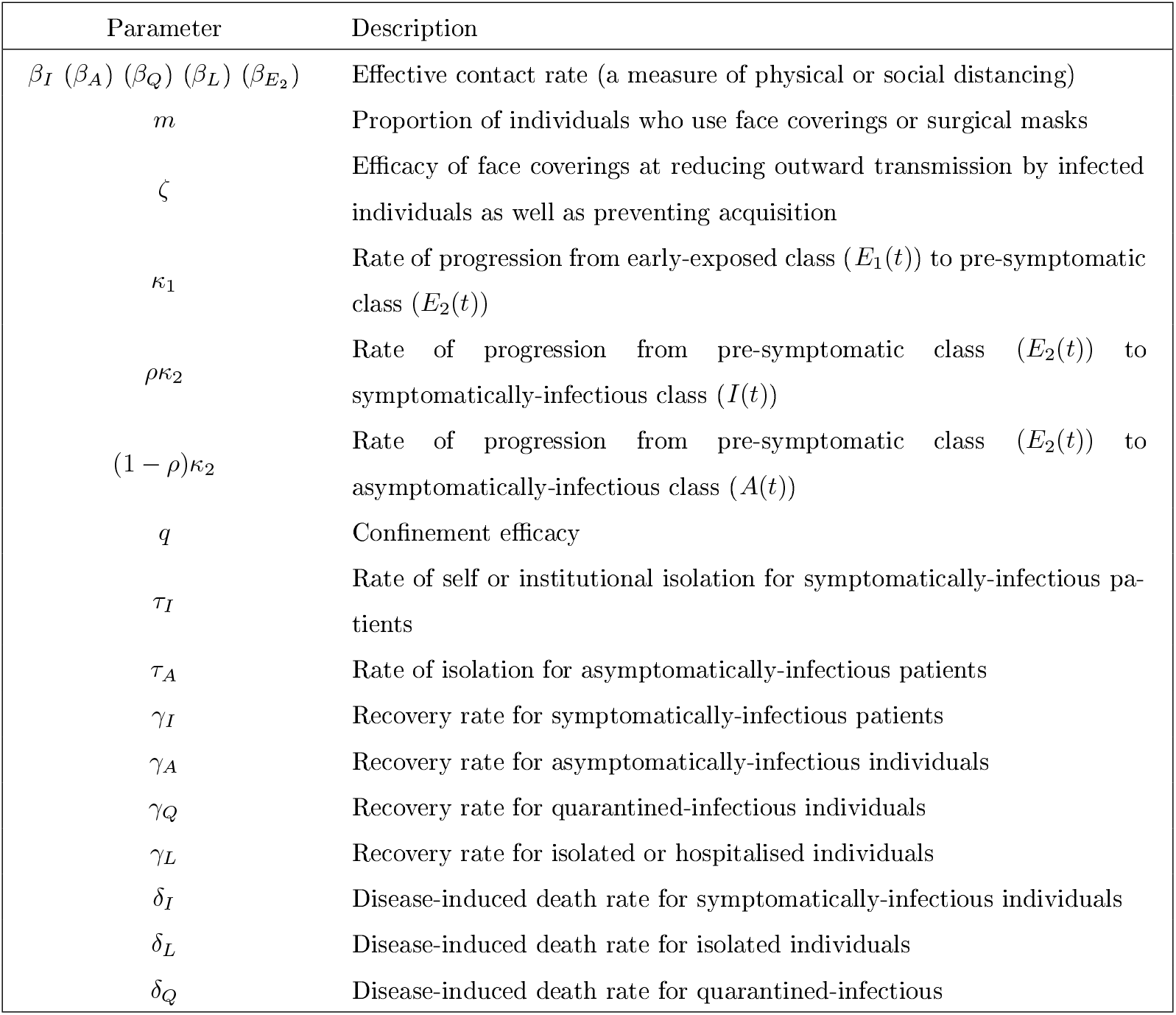
Model parameters and meaning

We set *x* = (*S, E*_1_, *E*_2_, *I, A, Q, L, R, D*)^*′*^ the vector of state variable, Let *f* : ℝ^9^ *→*ℝ^9^ the the right hand side of system (1), which is a continuously differentiable function on ℝ^9^. According to 23, Theorem III.10.VI, for any initial condition in Ω, a unique solution of (1) exists, at least locally, and remains in Ω for its maximal interval of existence [23, Theorem III.10.XVI]. Hence, model (1) is biologically well-defined.

### 2.2. Data sources

From the beginning of the COVID-19 outbreak, Center of Disease Control and Prevention (CDC) is providing authoritative and genuine data of daily confirmed COVID-19 cases. Daily confirmed data of five different countries named Argentina, Bangladesh, Brazil, Colombia and India have been compiled using that data repository. Johns Hopkins University Center for Systems Science and Engineering (JHU CSSE) is carefully maintaining the data repository supported by ESRI Living Atlas Team and the Johns Hopkins University Applied Physics Lab (JHU APL). The repository is really simple to use and publicly available [24].

## 3. Mathematical analysis

### 3.1. Asymptotic stability of disease-free equilibria

The disease-free equilibrium point denoted by *x*_0_ can be defined as follows:

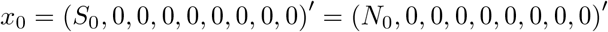

Using notations in 25, matrices *F* and *V* for the new infection terms and the remaining transfer terms are, respectively, given by

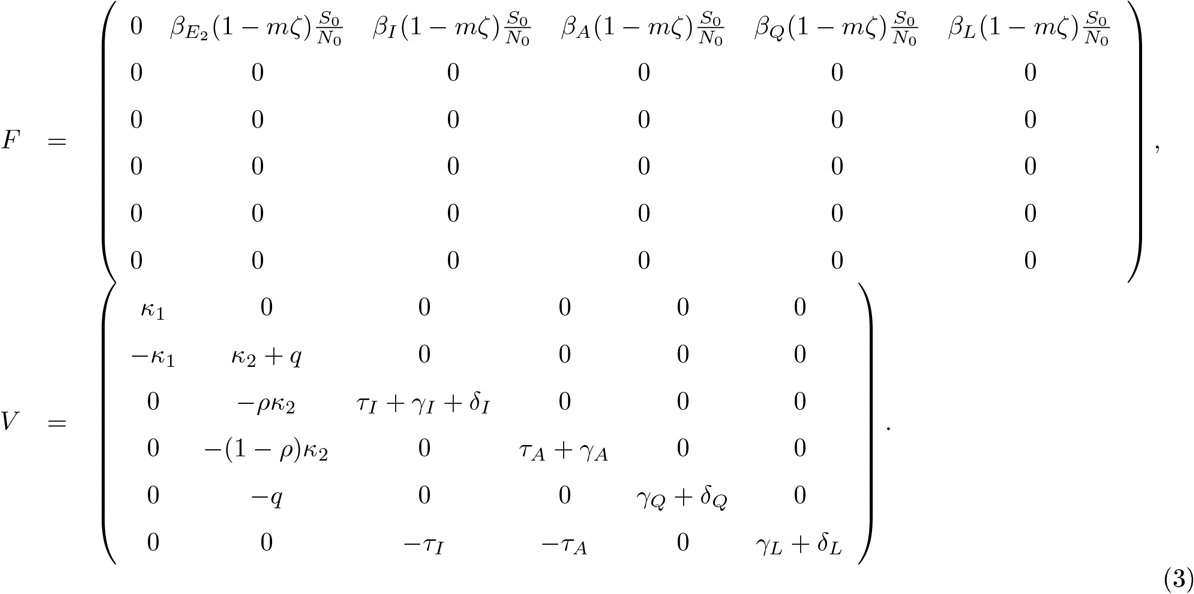

Then, the control reproduction ratio is defined, following [26, 25], as the spectral radius of the next generation matrix, *FV* ^*−*1^:

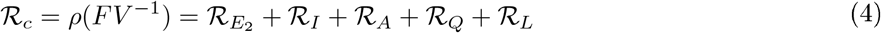

where,

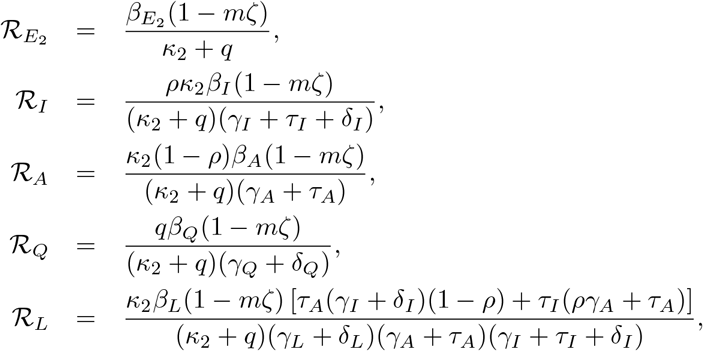

where *ρ*(·) represents the spectral radius operator.

The formula for control reproduction number has been formulated. Indeed, the insightful epidemic threshold, *ℛ*_*c*_ calculates the average number of new secondary COVID-19 cases generated by a COVID-19 positive individual in a population a portion susceptible people are using effective face coverings. Different non-pharmaceutical measures are acting as control measures which lead to bring down *ℛ*_*c*_ under unity 25. Hence, we claim the following result followed by a direct consequence of the next generation operator method 25, Theorem 2. where *ρ*(·) represents the spectral radius operator. The insightful epidemic threshold, *ℛ*_0_ calculates the average number of new secondary COVID-19 infections generated by an COVID-19 positive patient in a completely susceptible population. The control of COVID-19 pandemic passes by the application of some control measures which contribute to decrease until *ℛ*_0_ less than one 25. Hence we claim the following result.

#### Theorem 1.

The COVID-19 transmission dynamics is influenced by the basic reproduction number *ℛ*_0_ as follows:

1. If *ℛ*_0_ *<* 1, then a sufficiently small flow of infected individuals will not generate an outbreak of the COVID-19,i.e the disease-equilibrium *ε*_0_ is locally asymptotically stable on *ω*.
2. If *ℛ*_0_ *>* 1, then a sufficiently small flow of infected individuals will generate an outbreak of the COVID-19, and the disease-equilibrium *ε*_0_ is unstable.

#### Lemma 1.

If *ℛ*_*c*_ *<* 1, the disease-free equilibrium *x*_0_ is locally asymptotically stable and unstable if *ℛ*_*c*_ *>* 1.

#### Remark 2.

Lemma 1 implies that if *ℛ*_*c*_ *<* 1, then a sufficiently small flow of infected individuals will not generate an outbreak of COVID-19, whereas for *ℛ*_*c*_ *>* 1, epidemic curve reaches a peak by growing exponentially and then decreases to zero as *t → ∞*.

The better control of the COVID-19 can be established by the fact that the DFE *x*_0_ is globally asymptotically stable (GAS). In this context, we claim the following result.

#### Theorem 3.

if *ℛ*_*c*_ *<* 1, then the manifold, *𝒲*, of disease-free equilibrium points of the model (1) is GAS in *𝒟*.

In the absence of use of face coverings, i.e. *m* = 0, *ℛ*_*c*_ converges to the basic reproduction number, *ℛ*_0_. Now, we will study the global stability of the disease-free equilibrium whenever the basic reproduction number is less than one (*ℛ*_*c*_ *<* 1). For this, we use the following Lyapunov function

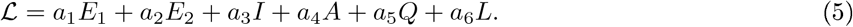

By deriving this function along the trajectories of the system (1), we obtain

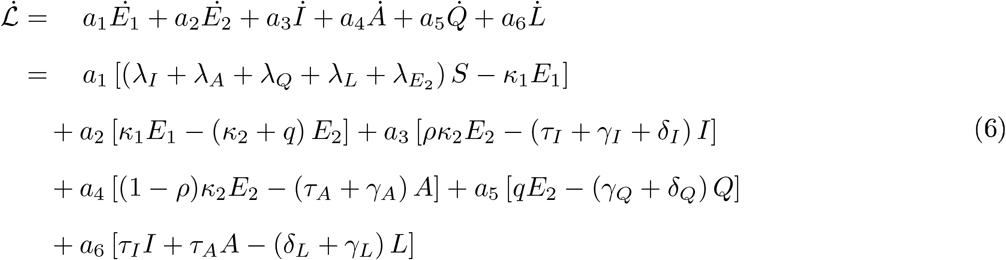

Since 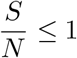, we obtain

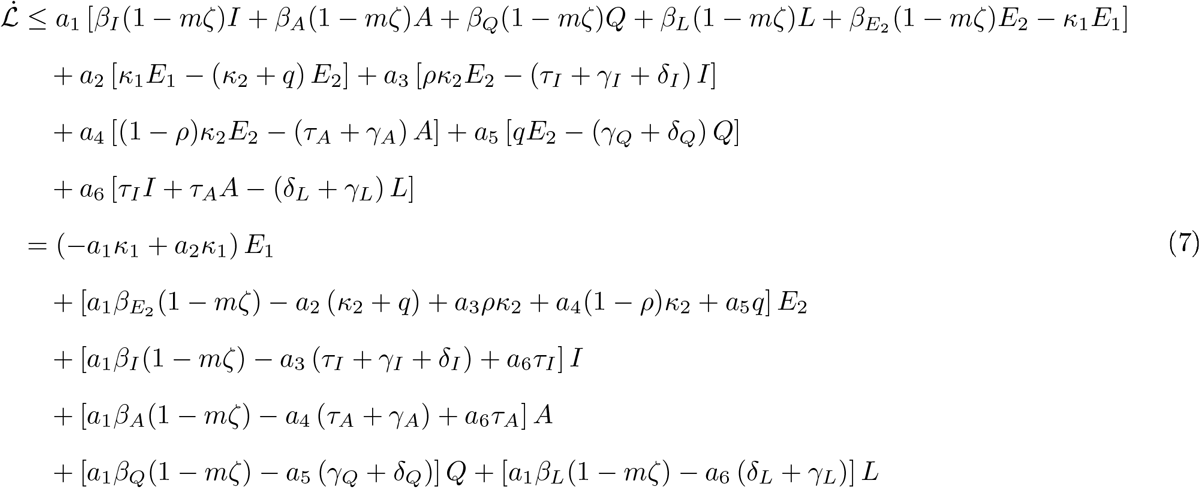

We choose *a*_*i*_, *i* = 1, 2, …, 6, such that coefficients of *E*_1_, *I, A, Q*, and *L* become zero. That is

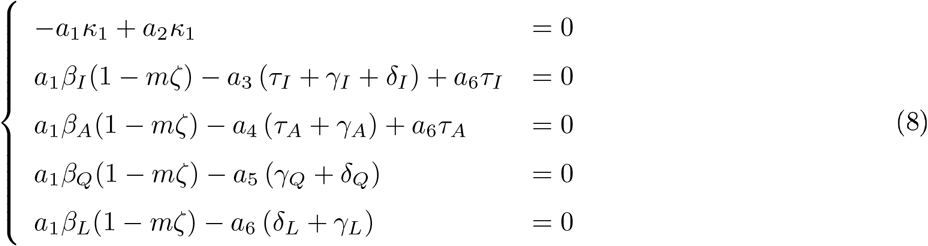

which the non-zero solution is given by

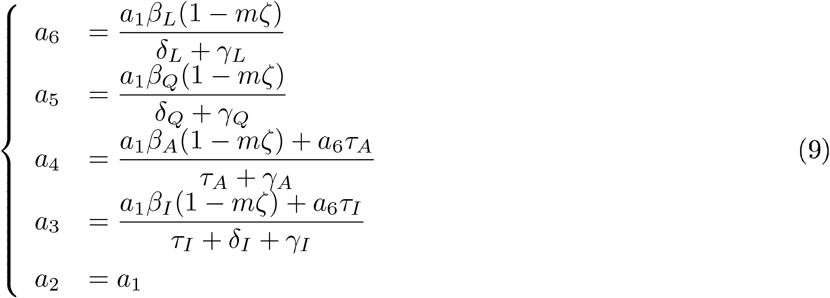

Plugging (9) into (7) gives

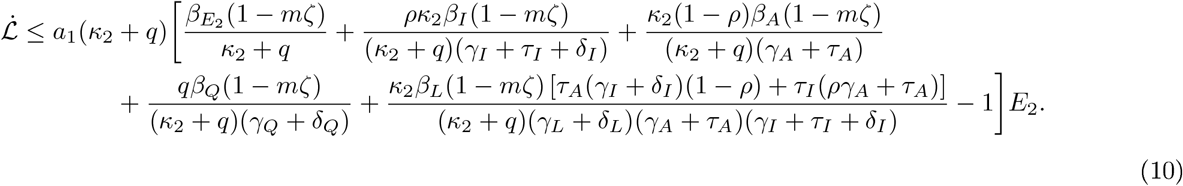

Setting 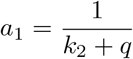, we finally obtain

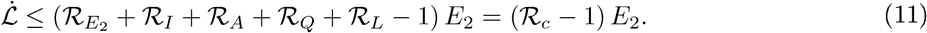

From (11), it follows that 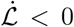 if *ℛ*_*c*_ *<* 1, and 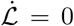 if and only if *E*_1_ = *E*_2_ = *I* = *A* = *Q* = *L* = 0. Therefore, *ℒ* is a Lyapunov function for system (1). Moreover, the maximal invariant set contained in 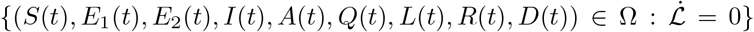 is the continuum of the disease-free equilibrium (*ε*_0_). Thus, from Lyapunov theory, we deduce that the disease-free equilibrium *ε*_0_ is GAS if *ℛ*_*c*_ *<* 1. Hence, it follows, by the LaSalle’s Invariance Principal, that the continuum of disease-free equilibria of the model (1) is a stable global attractor in Ω whenever *ℛ*_*c*_ ≤ 1 The previous analysis can be summarize as follows:

#### Theorem 4.

If *ℛ*_*c*_ ≤ 1, then the disease-free equilibrium *ε*_0_ is globally asymptotically stable on Ω.

## 4. Model calibration and forecasting

The model (1) calibration has been performed using a newly developed optimization algorithm based on trust-region-reflective (TRR) algorithm, which can be regarded as an evolution of Levenberg-Marquardt algorithm 5. This robust optimization procedure can be used effectively for solving nonlinear least-squares problems. This algorithm has been implemented using the **lsqcurvefit** function, which is available in the Optimization Toolbox in MATLAB. Necessary model parameters have been estimated using this optimization technique. Daily infected cases data have been collected from a trusted data repository, which is available online. A 7-day moving average of the daily reported cases has been used for our model calibration due to moderate volatile nature of real data. It has been observed that the number of daily testing in Argentina, Bangladesh, Brazil, Colombia and India have been really inconsistent. With an aim to capture the real outbreak scenario, the 7-day moving average has been used in this regard.

### 4.1. Argentin

On March 3, 2020, Argentina registered its first COVID-19 case. As a consequence, government officials immediately deploy strict lockdown, quarantine and isolation measures to curb the spread of the novel coronavirus in the community. Due to unbearable economic crisis, inhabitants of the country have already started violating staying at home and quarantine orders. The country could face a second wave of infection in near future in the absence of non-pharmaceutical intervention measures. The robustness of our model fitting performance has been illustrated in Figs. 2 and 3 as of November 11, 2020. Moreover, projection results from mid November to early April for daily and cumulative cases in Argentina have been depicted in Figs. 4 and 5. According to our projection results, the daily could reach upto 2450*K* cases by the end of March 2021, if the current trend holds. Model parameters presented in the Table 2 have been calibrated using observed real data from March 4 to November 11, 2020.

**Table 2:** Calibrated parameters of the proposed model (1) using trust-region-re ective algorithm and daily COVID-19 cases data of Argentina

| Parameter | Range (Unit) | Baseline<br>value | TRR output | Reference |
| --- | --- | --- | --- | --- |
| $\beta_I$ | 0.1–1.5 $day^{-1}$ | 0.55 | 0.3 | [27, 5] |
| $\beta_A$ | 0.1–0.9 $day^{-1}$ | 0.3 | 0.15 | [27, 5] |
| $\beta_Q$ | 0.1–0.9 $day^{-1}$ | 0.5 | 0.3 | [27, 5] |
| $\beta_L$ | 0.1–0.9 $day^{-1}$ | 0.3 | 0.35 | [5, 6] |
| $\beta_{E_2}$ | 0.05–0.3 $day^{-1}$ | 0.3 | 0.1 | [6] |
| $m$ | 0.01–0.3 (dimensionless) | 0.1 | 0.15 | [6] |
| $\zeta$ | 0.5 (dimensionless) | 0.5 | 0.5 | [6] |
| $\kappa_1$ | $\frac{1}{4} day^{-1}$ | $\frac{1}{4}$ | $\frac{1}{4}$ | [3, 6] |
| $\kappa_2$ | 1 $day^{-1}$ | 1 | 1 | [3] |
| $q$ | 0.1–0.6 $day^{-1}$ | 0.3 | 0.25 | Estimated |
| $\rho$ | 0.6–0.7 (dimensionless) | 0.65 | 0.65 | [6] |
| $\tau_I$ | $\frac{1}{14} - \frac{1}{5} day^{-1}$ | 1/10 | 1/10 | [5, 6] |
| $\tau_A$ | $\frac{1}{14} - \frac{1}{5} day^{-1}$ | 1/10 | 1/10 | [5, 6] |
| $\gamma_I$ | $\frac{1}{14} - \frac{1}{7} day^{-1}$ | 1/7 | 0.143 | [28, 29] |
| $\gamma_A$ | $\frac{1}{10} - \frac{1}{7} day^{-1}$ | 1/7 | 0.143 | [28, 29] |
| $\gamma_Q$ | $\frac{1}{21} - \frac{1}{10} day^{-1}$ | 1/14 | 0.071 | [28, 29] |
| $\gamma_L$ | $\frac{1}{21} - \frac{1}{10} day^{-1}$ | 1/14 | 0.071 | [28, 29] |
| $\delta_I$ | 0.0001–0.01 $day^{-1}$ | 0.001 | 0.00033 | [2, 5] |
| $\delta_L$ | 0.0001–0.01 $day^{-1}$ | 0.001 | 0.001 | [2, 5] |
| $\delta_Q$ | 0.0001–0.01 $day^{-1}$ | 0.001 | 0.00033 | [2, 5] |

**Figure 2:**
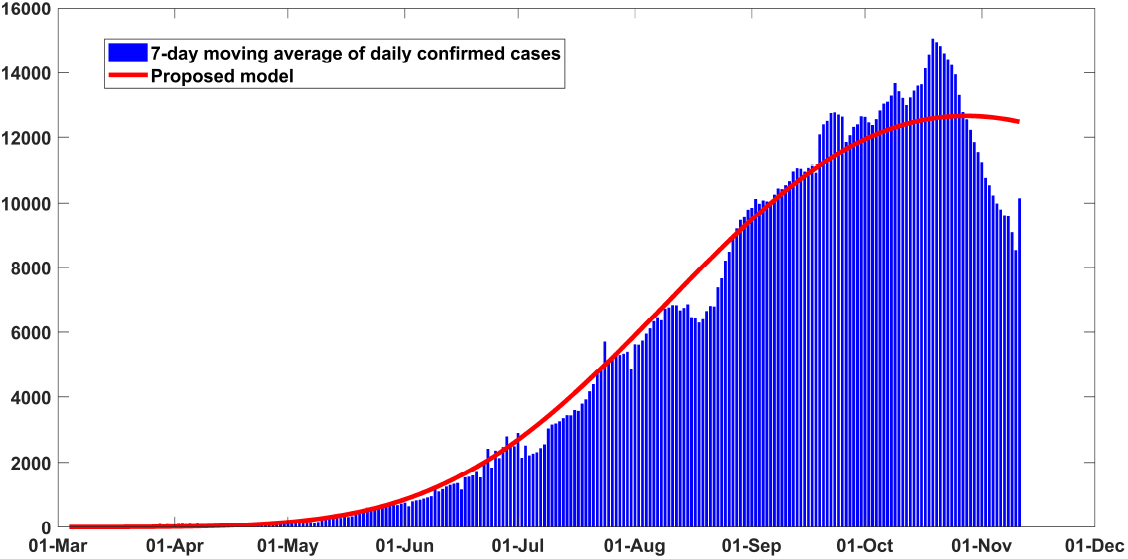
Fitting performance of the model for daily infected cases in Argentina from March 04 to November 11, 2020.

**Figure 3:**
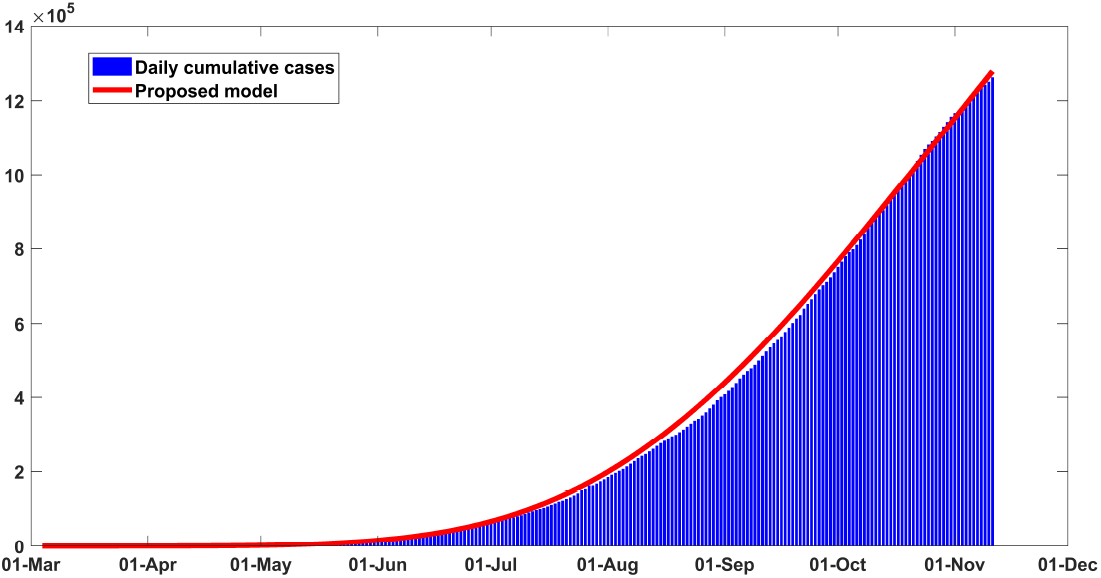
Fitting performance of the model for cumulative infected cases in Argentina from March 04 to November 11, 2020.

**Figure 4:**
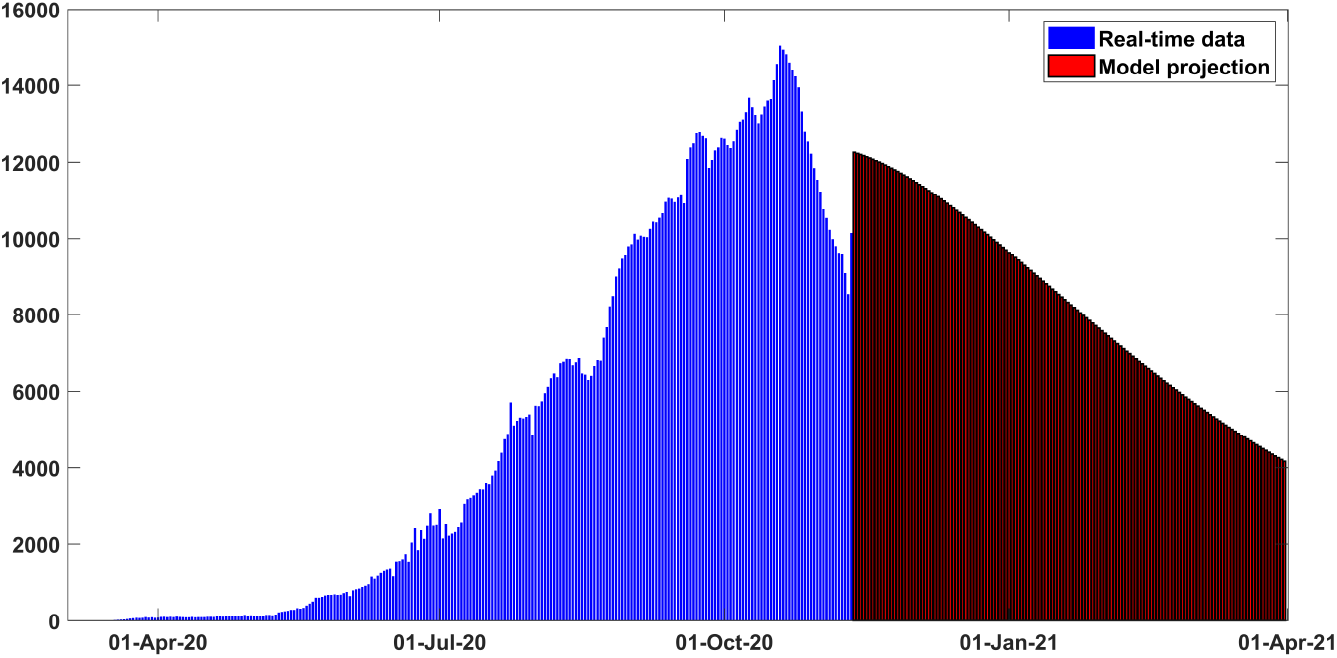
Projection results for daily new confirmed cases for Argentina from early March to late March 2021.

**Figure 5:**
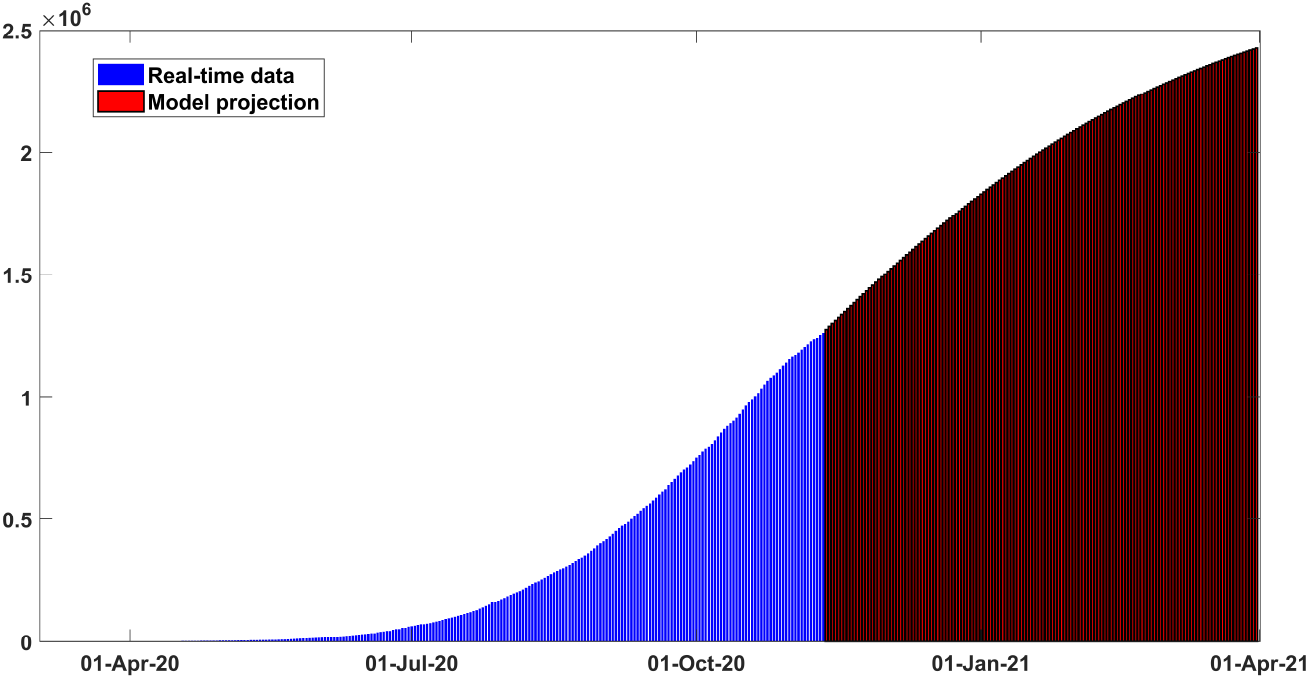
Projection results for cumulative cases for Argentina early March to late March 2021.

As we can see the results from the proposed model responses complement the real data very well. The control reproduction number (*ℛ*_*c*_) is estimated to be *∼* 1.37 (95% *CI* : 1.03 *−* 1.59) as of November 11 and prior established findings 1 *−* 5 for COVID-19 really match well with the estimation [30, 5]. The tally of cumulative infected cases is projected to reach 2450*K* and country’s death toll could mount to 52.6*K* by the end of March 2021. Table 2 illustrates model calibration results and baseline parameter values.

### 4.2. Bangladesh

Due to prolonged lockdown measures and severe economic recession, inhabitants of Bangladesh have started violating safety measures like wearing face coverings and maintaining physical distancing. Figs. 6 and 7 illustrate the model fitting performance with observed data from early March to mid November for Bangladesh. The estimated error is found to be hovering around 10% for daily new cases. The actual outbreak scenario in Bangladesh is still a puzzle to be solved for the health officials due to scant COVID-19 testing program. The control reproduction number (*ℛ*_*c*_) is estimated to be *∼* 1.17 (95% *CI* : 0.95 *−* 1.39) as of November 11 and prior established findings for this metric go well with the estimation [30, 5]. The tally of cumulative infected cases is projected to reach 436*K* around March 31, 2021 and the estimated total death cases could reach 11, 400 by the end of March, 2021. Table 3 illustrates the key features used to calibrate this scenario, which have been justified in prior clinical studies and relevant literature.

**Table 3:** Calibrated parameters of the proposed model (1) using trust-region-re ective algorithm and daily COVID-19 cases data of Bangladesh

| Parameter | Range (Unit) | Baseline<br>value | TRR output | Reference |
| --- | --- | --- | --- | --- |
| $\beta_I$ | 0.1–1.5 $day^{-1}$ | 0.55 | 0.15 | [27, 5] |
| $\beta_A$ | 0.1–0.9 $day^{-1}$ | 0.3 | 0.1 | [27, 5] |
| $\beta_Q$ | 0.1–0.9 $day^{-1}$ | 0.5 | 0.1 | [27, 5] |
| $\beta_L$ | 0.1–0.9 $day^{-1}$ | 0.3 | 0.12 | [5, 6] |
| $\beta_{E_2}$ | 0.05–0.3 $day^{-1}$ | 0.3 | 0.11 | [6] |
| $m$ | 0.01–0.3 (dimension-<br>less) | 0.1 | 0.3 | [6] |
| $\zeta$ | 0.5 (dimensionless) | 0.5 | 0.5 | [6] |
| $\kappa_1$ | $\frac{1}{4} day^{-1}$ | $\frac{1}{4}$ | $\frac{1}{4}$ | [3, 6] |
| $\kappa_2$ | 1 $day^{-1}$ | 1 | 1 | [3] |
| $q$ | 0.1–0.6 $day^{-1}$ | 0.3 | 0.47 | Estimated |
| $\rho$ | 0.6–0.7 (dimension-<br>less) | 0.65 | 0.65 | [6] |
| $\tau_I$ | $\frac{1}{14} - \frac{1}{5} day^{-1}$ | 1/10 | 1/8 | [5, 6] |
| $\tau_A$ | $\frac{1}{14} - \frac{1}{5} day^{-1}$ | 1/10 | 1/8 | [5, 6] |
| $\gamma_I$ | $\frac{1}{14} - \frac{1}{7} day^{-1}$ | 1/7 | 1/12 | [28, 29] |
| $\gamma_A$ | $\frac{1}{10} - \frac{1}{7} day^{-1}$ | 1/7 | 1/10 | [28, 29] |
| $\gamma_Q$ | $\frac{1}{21} - \frac{1}{10} day^{-1}$ | 1/21 | 0.071 | [28, 29] |
| $\gamma_L$ | $\frac{1}{21} - \frac{1}{10} day^{-1}$ | 1/21 | 0.071 | [28, 29] |
| $\delta_I$ | 0.0001–0.01 $day^{-1}$ | 0.001 | 0.0004 | [2, 5] |
| $\delta_L$ | 0.0001–0.01 $day^{-1}$ | 0.001 | 0.0009 | [2, 5] |
| $\delta_Q$ | 0.0001–0.01 $day^{-1}$ | 0.001 | 0.0007 | [2, 5] |

**Figure 6:**
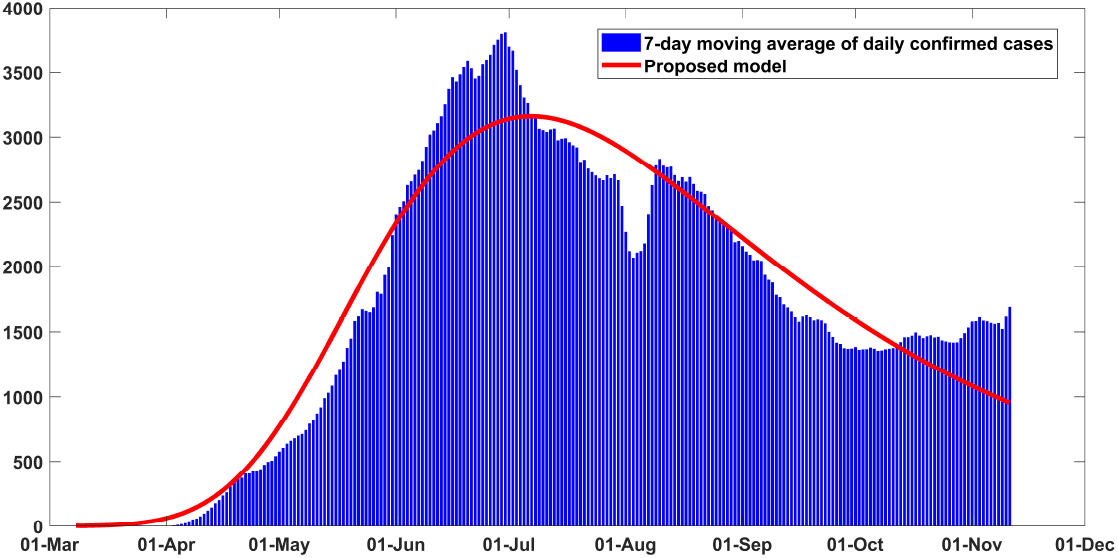
Fitting performance of the model for daily infected cases in Bangladesh from March 08 to November 11, 2020.

**Figure 7:**
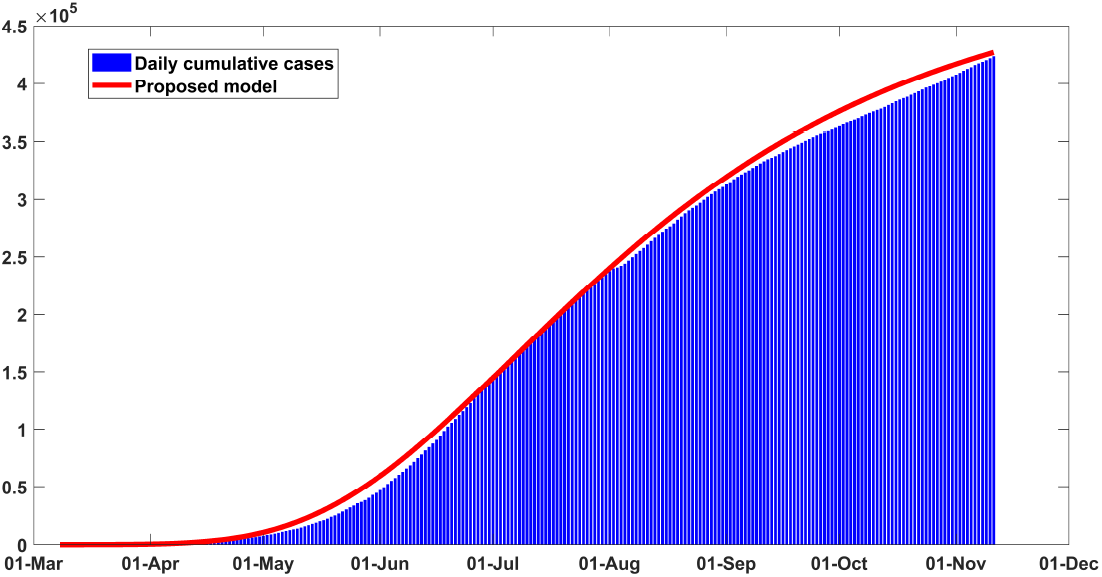
Fitting performance of the model for cumulative infected cases in Bangladesh from March 08 to November 11, 2020.

### 4.3. Brazil

COVID-19 pandemic emerged in Latin America a bit later than other continents and Brazil is among the hardest-hit countries in the world. When late-stage clinical trials of Chinese-developed CoronaVac vaccine are going on in Brazil, the inhabitants of Brazil are witnessing unprecedented spikes in new COVID-19 cases.

As of November 15, 2020, Brazil reported 5,863,093 cases and related 165,811 deaths. The model fitting performance for Brazil from late February to mid November are illustrated in Figs. 10 and 11. Historical data from late February to November 11, 2020 have been considered to calibrate the model parameters. It is clearly visible in the figures that the responses from the proposed model fit the real data very well. The control reproduction number is estimated about *∼* 2.12 as of November 11, which is in between the observed findings for COVID-19 30, 5. Fig. 13 illustrates that the tally of total infected cases could mount to 6368.6*K* by the end of March 2021 if current trend is held, and death toll could surpass 215*K* within this time period. Table 4 depicts the key features used to calibrate this scenario.

**Table 4:**
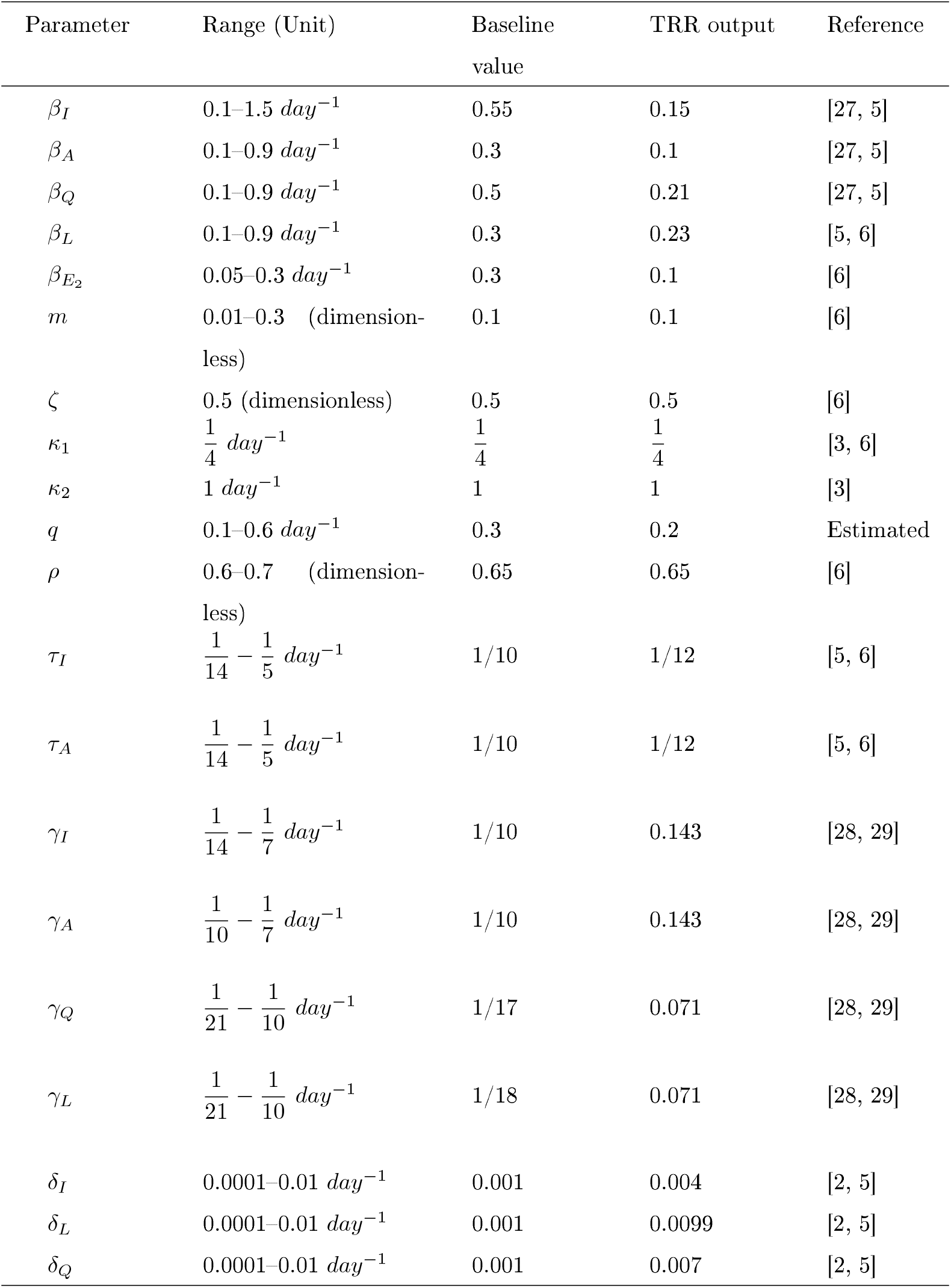
Calibrated parameters of the proposed model (1) using trust-region-re ective algorithm and daily COVID-19 cases data of Brazil

| Parameter | Range (Unit) | Baseline<br>value | TRR output | Reference |
| --- | --- | --- | --- | --- |
| $\beta_I$ | 0.1–1.5 $day^{-1}$ | 0.55 | 0.15 | [27, 5] |
| $\beta_A$ | 0.1–0.9 $day^{-1}$ | 0.3 | 0.1 | [27, 5] |
| $\beta_Q$ | 0.1–0.9 $day^{-1}$ | 0.5 | 0.21 | [27, 5] |
| $\beta_L$ | 0.1–0.9 $day^{-1}$ | 0.3 | 0.23 | [5, 6] |
| $\beta_{E_2}$ | 0.05–0.3 $day^{-1}$ | 0.3 | 0.1 | [6] |
| $m$ | 0.01–0.3 (dimension-<br>less) | 0.1 | 0.1 | [6] |
| $\zeta$ | 0.5 (dimensionless) | 0.5 | 0.5 | [6] |
| $\kappa_1$ | $\frac{1}{4} day^{-1}$ | $\frac{1}{4}$ | $\frac{1}{4}$ | [3, 6] |
| $\kappa_2$ | 1 $day^{-1}$ | 1 | 1 | [3] |
| $q$ | 0.1–0.6 $day^{-1}$ | 0.3 | 0.2 | Estimated |
| $\rho$ | 0.6–0.7 (dimension-<br>less) | 0.65 | 0.65 | [6] |
| $\tau_I$ | $\frac{1}{14} - \frac{1}{5} day^{-1}$ | 1/10 | 1/12 | [5, 6] |
| $\tau_A$ | $\frac{1}{14} - \frac{1}{5} day^{-1}$ | 1/10 | 1/12 | [5, 6] |
| $\gamma_I$ | $\frac{1}{14} - \frac{1}{7} day^{-1}$ | 1/10 | 0.143 | [28, 29] |
| $\gamma_A$ | $\frac{1}{10} - \frac{1}{7} day^{-1}$ | 1/10 | 0.143 | [28, 29] |
| $\gamma_Q$ | $\frac{1}{21} - \frac{1}{10} day^{-1}$ | 1/17 | 0.071 | [28, 29] |
| $\gamma_L$ | $\frac{1}{21} - \frac{1}{10} day^{-1}$ | 1/18 | 0.071 | [28, 29] |
| $\delta_I$ | 0.0001–0.01 $day^{-1}$ | 0.001 | 0.004 | [2, 5] |
| $\delta_L$ | 0.0001–0.01 $day^{-1}$ | 0.001 | 0.0099 | [2, 5] |
| $\delta_Q$ | 0.0001–0.01 $day^{-1}$ | 0.001 | 0.007 | [2, 5] |

**Figure 8:**
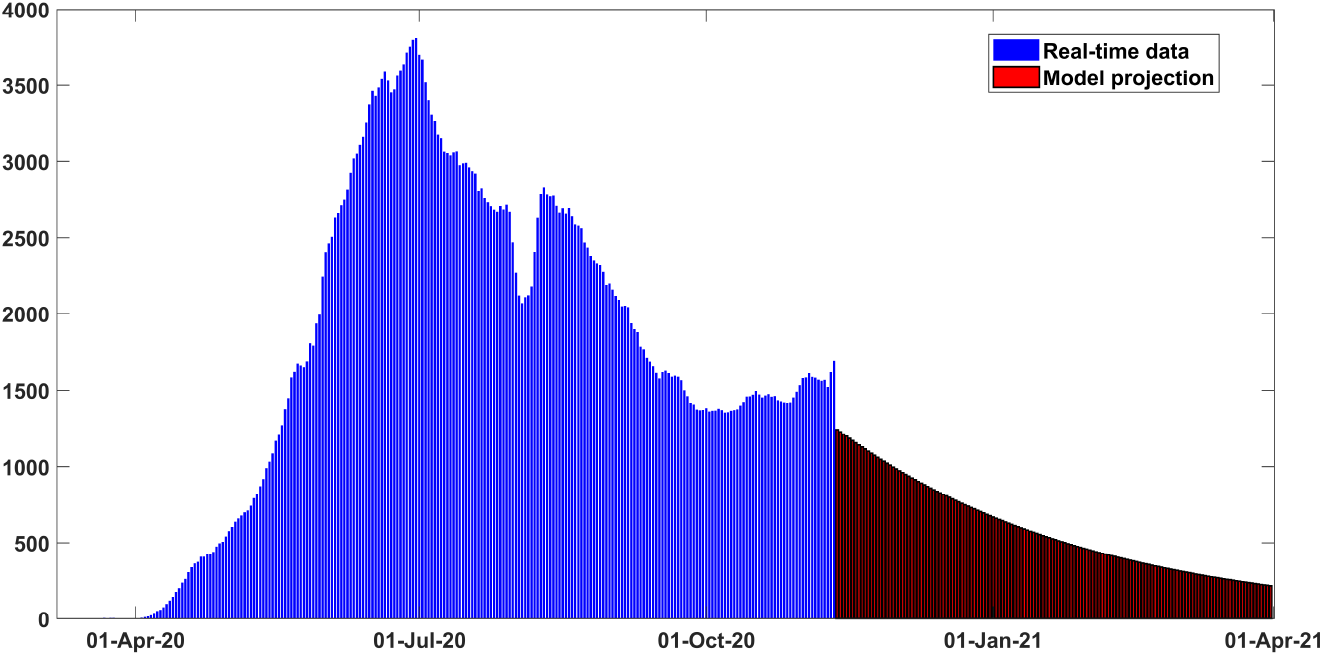
Projection results for daily new confirmed cases for Bangladesh from early March to late March 2021.

**Figure 9:**
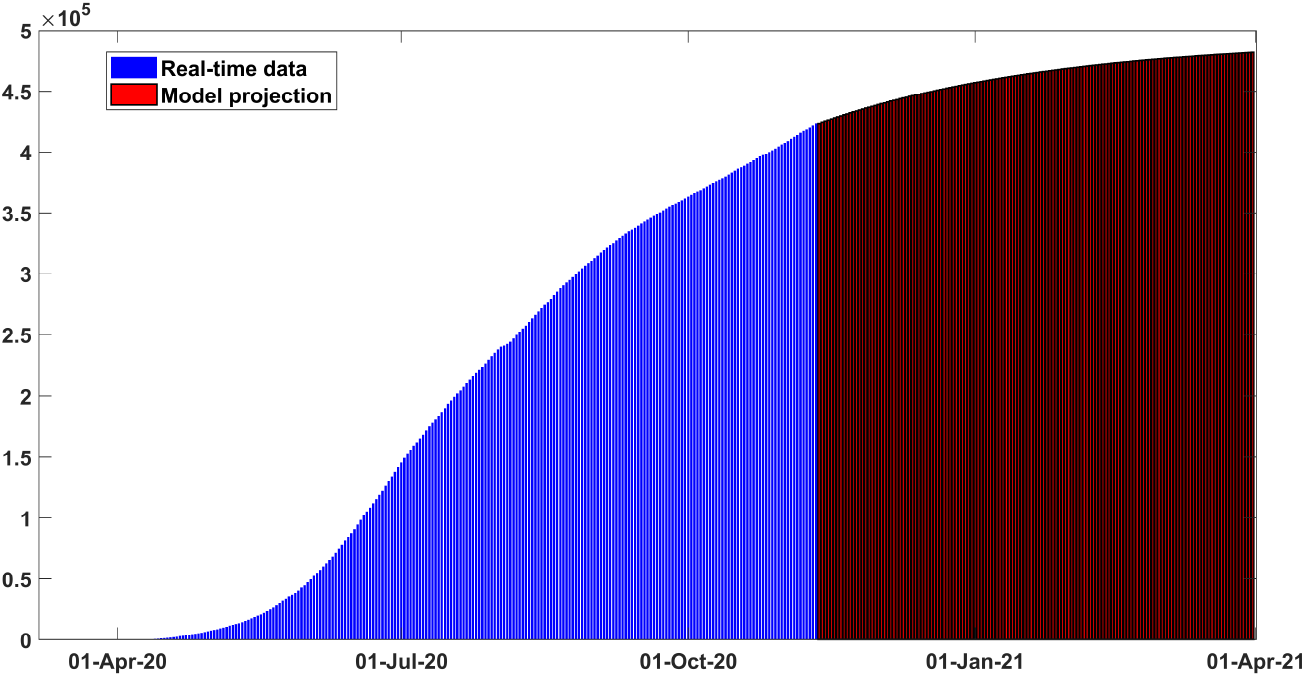
Projection results for cumulative cases for Bangladesh early March to late March 2021.

**Figure 10:**
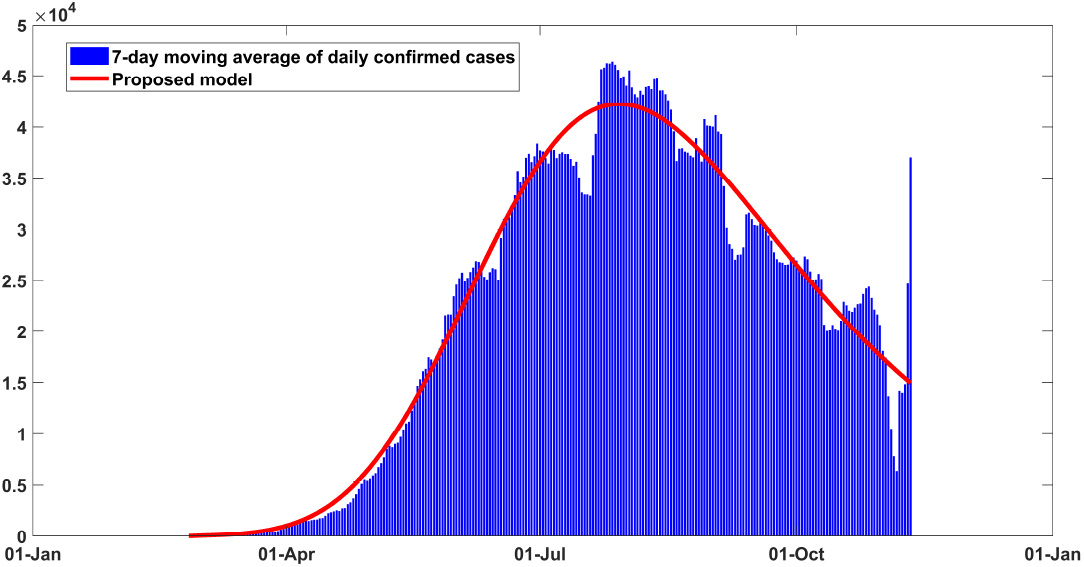
Fitting performance of the model for daily infected cases in Brazil from February 25 to November 11, 2020.

**Figure 11:**
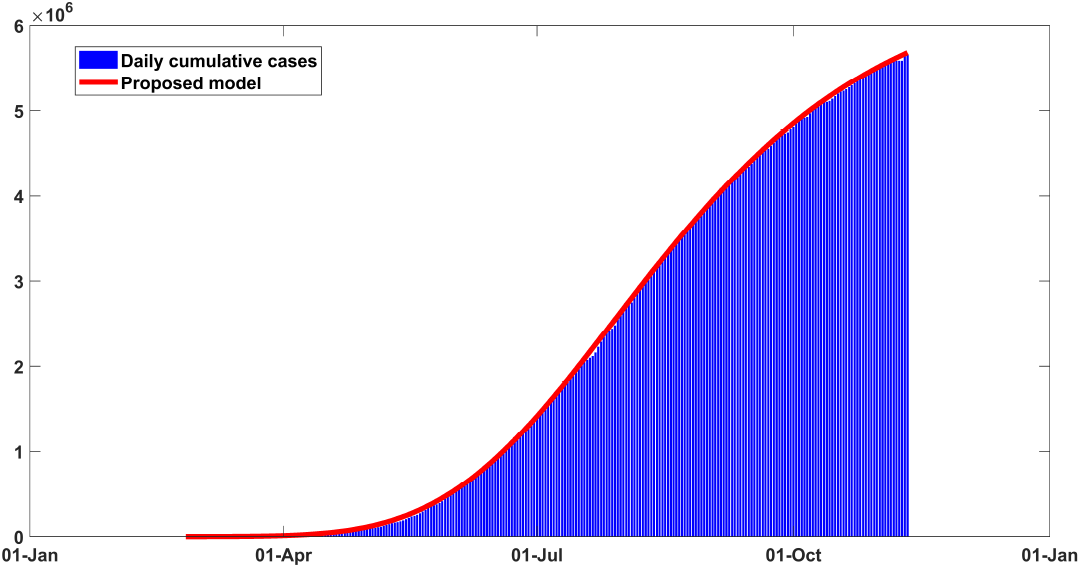
Fitting performance of the model for cumulative infected cases in Brazil from February 25 to November 11, 2020.

**Figure 12:**
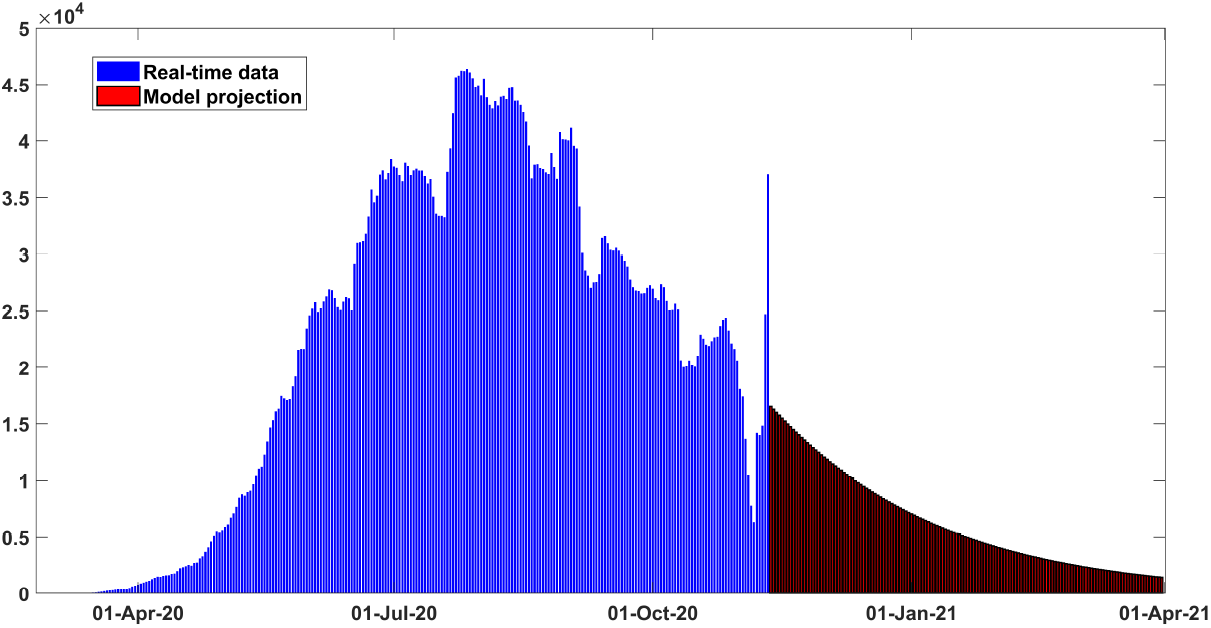
Projection results for daily new confirmed cases for Brazil from early March to late March 2021.

**Figure 13:**
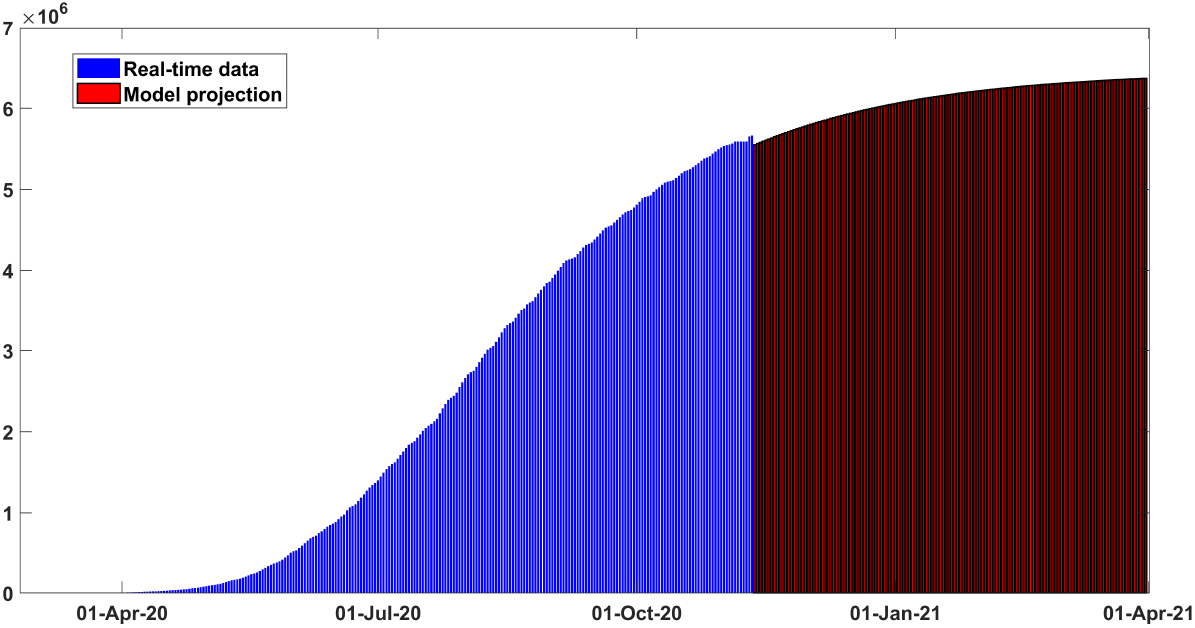
Projection results for cumulative cases for Brazil early March to late March 2021.

### 4.4. Colombia

Colombia is one of the hardest-hit countries in Latin America which has already surpassed one million confirmed coronavirus cases. As of November 15, 2020, Colombia has seen 1,198,746 confirmed cases of COVID-19 and the country’s death toll climbed to 34,031. Our model fitting performance with the historical data is quite outstanding depicted in Figs. 14 and 15. According to our projection results illustrated in Figs 16 and 17, the tally of cumulative cases could reach approximately 1730*K* cases and the number of daily cases could hover around 2000 cases by the end of March 2021. The control reproduction number (*ℛ*_┘_) is estimated to be *∼* 1.19 as of November 11, 2020. According to our analysis, the epidemic threshold (*ℛ*_┘_) could be brought under unity by the end of December 2020 by following strict distance maintaining protocol and mass-level usage of efficacious face coverings.

**Figure 14:**
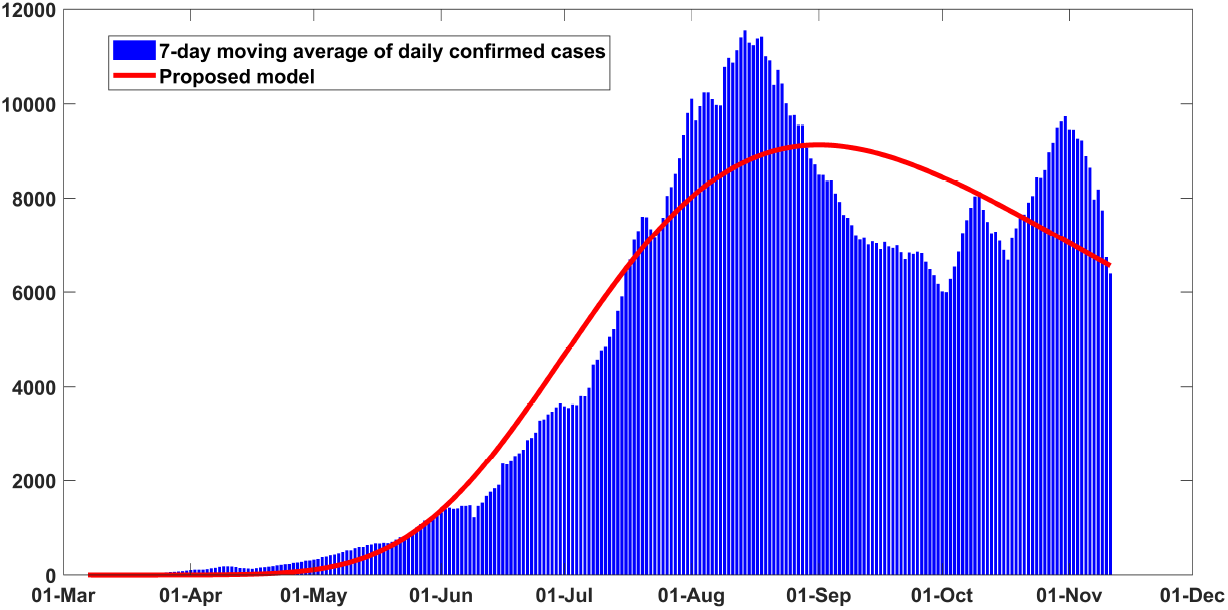
Fitting performance of the model for daily infected cases in Colombia from March 06 to November 11, 2020.

**Figure 15:**
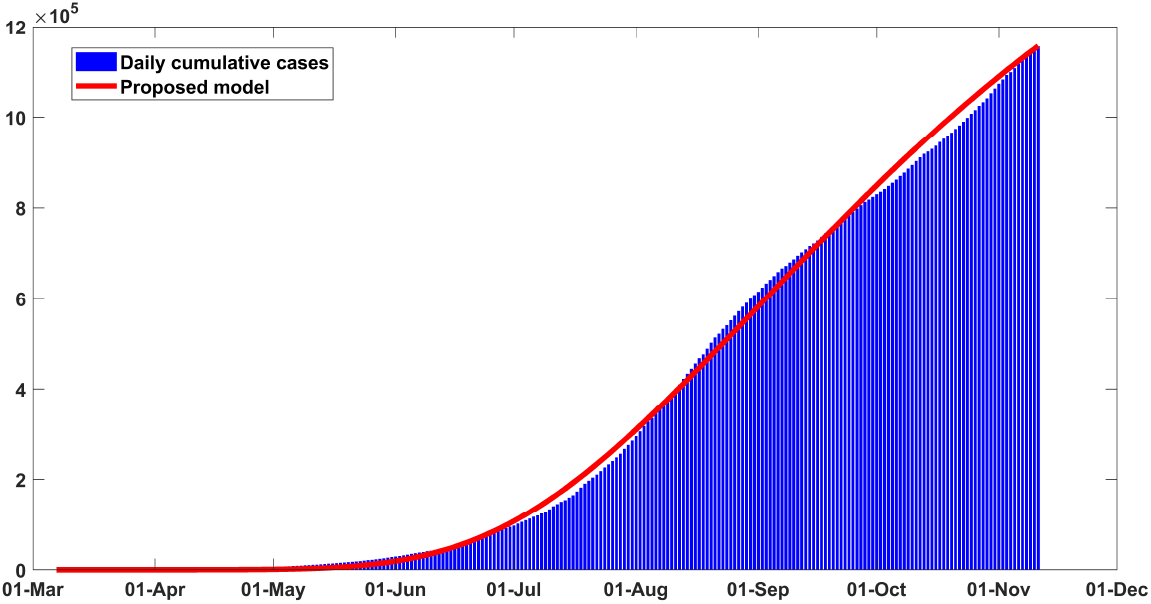
Fitting performance of the model for cumulative infected cases in Colombia from March 06 to November 11, 2020.

**Figure 16:**
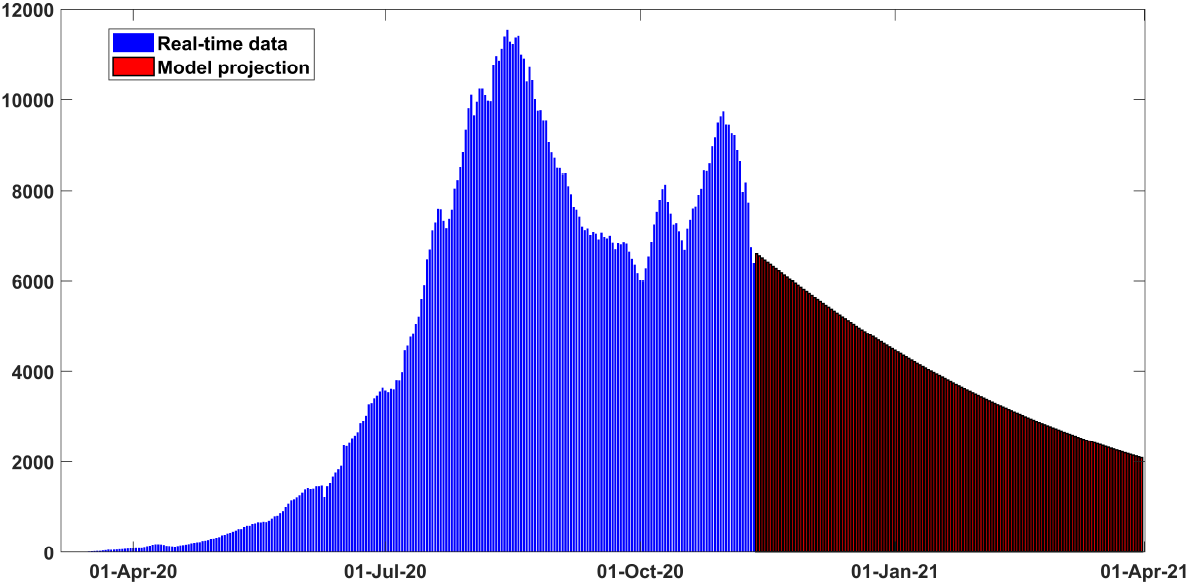
Projection results for daily new confirmed cases for Colombia from early March to late March 2021.

**Figure 17:**
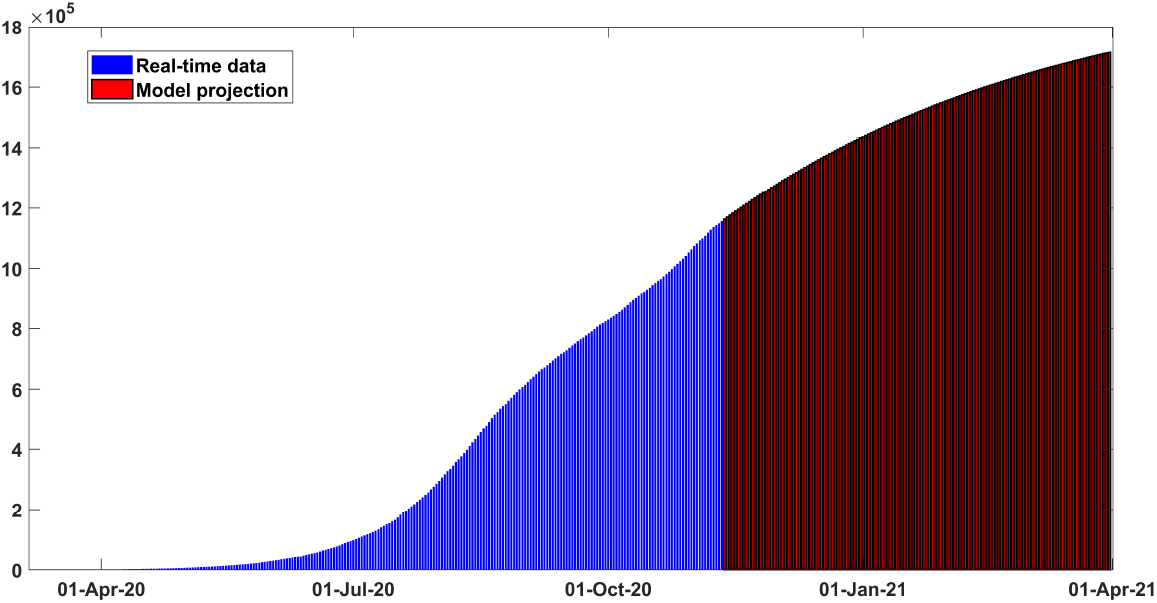
Projection results for cumulative cases for Colombia early March to late March 2021.

### 4.5. India

When India is celebrating a busy festival season, the tally of fresh COVID-19 cases continued to rise. Relaxation in protective and social-distancing measures could result in a significant upsurge in daily cases in upcoming days. Fig. 18 illustrates the fact that India is witnessing a downward trend after having peak. According to our projection results, India could reel under the second wave of infection unless non-pharmaceutical interventions strategies are followed comprehensively. As of November 15, the country’s caseload now stands at 8,850,338 and it’s death toll has mounted to 130,187. Figs. 18 and 19 illustrate the fitting performance of our proposed model for India from late January to mid November. Historical data from January 30 to November 11 have been considered to calibrate the model parameters. As we can see from the figures, model-fitting is exceptionally well for the historical observed data. Based on our projection results from Fig. 20, the number of daily cases could be brought under 1000 cases if mass-level efficacious face coverings is strictly maintained. The control reproduction number *ℛ*_*c*_ is estimated to be *∼* 1.41 as of November 11, which complements the prior studied observations [30, 5]. The tally of cumulative infected cases is projected to reach 11259*K* by the end of March 2021 if current trend continues. In addition, country’s death toll could mount to 187*K* over the period. Tab. 6 illustrates the key features used to calibrate this scenario.

**Figure 18:**
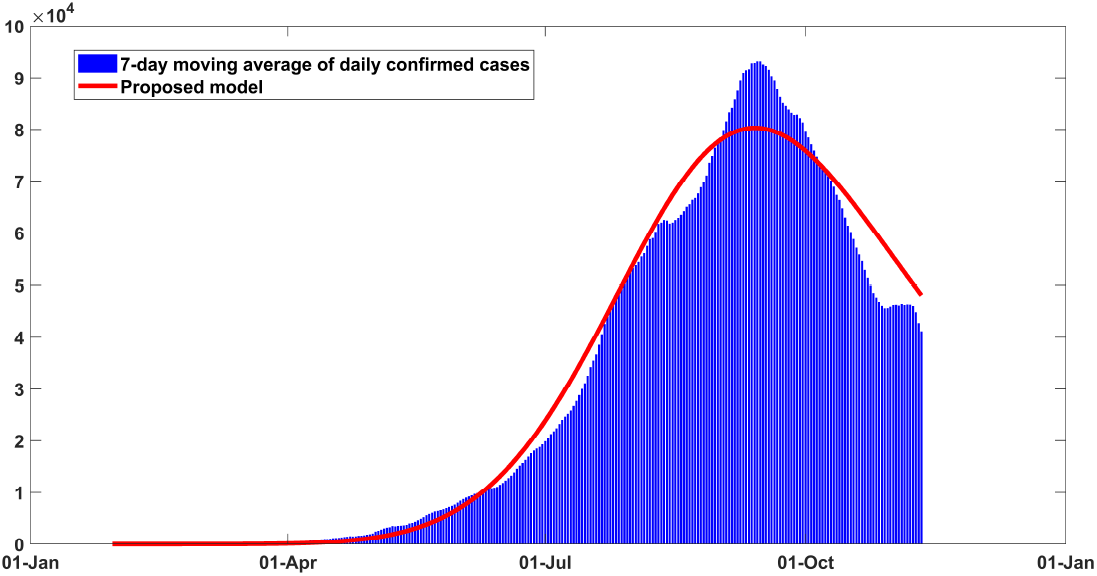
Fitting performance of the model for daily infected cases in India from January 30 to November 11, 2020.

**Figure 19:**
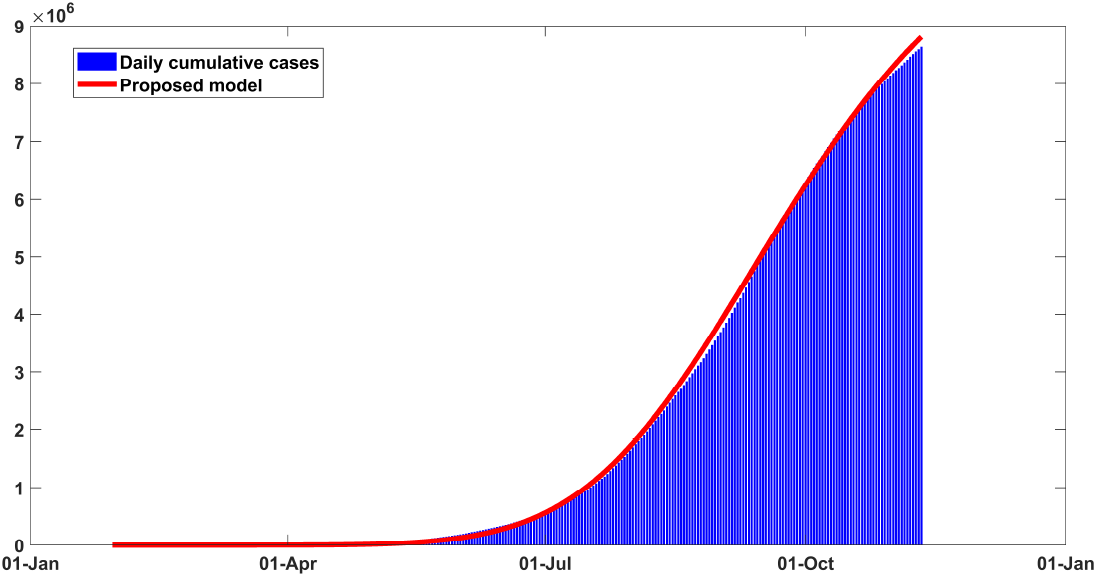
Fitting performance of the model for cumulative infected cases in India from January 30 to November 11, 2020.

**Figure 20:**
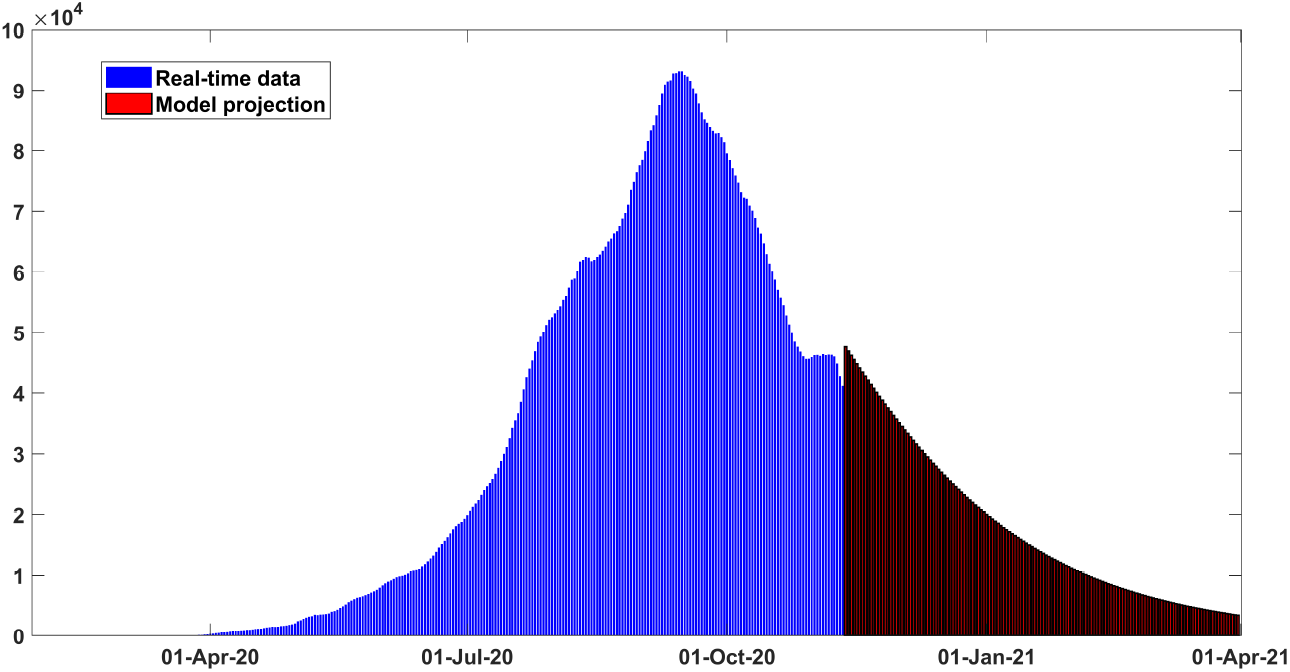
Projection results for daily new confirmed cases for India from late January 2020 to late March 2021.

## 5. Global sensitivity analysis

With an aim to quantify the most dominant mechanisms in the proposed model, a renowned global sensitivity analysis approach Partial Rank Correlation Coefficient (PRCC) method has been carried out. This method can provide considerable insights about the relationship between model responses (state variables) and model parameters (sampled by Latin Hypercube Sampling method) in an outbreak setting [31]. PRCC values are generally bounded between -1 and 1. A monotonic relationship between the model input parameters and the model outputs is generally assumed in this method. Apart from qualitative relationship, precise quantitative impact of different parameters on model responses can be determined by calculating the PRCC values. Uniform distribution of all model parameters have been considered to generate LHS matrices. A positive PRCC value depicts that the model output can be increased by increasing the respective model input parameter. Similarly, model output can be decreased by forcing down the corresponding input parameter. In addition, a negative PRCC value indicates a negative correlation between the model input and output.

When it comes to analyse a complex model, it often get really challenging to control the parameters. In this context, sensitivity analysis can give considerable insights regarding the quantitative relationship between model responses and model input parameters. However, it is really challenging for complex models to determine the qualitative and quantitative relationship with sufficient accuracy. As we can see from Figure 22, nearly the same qualitative relationship has been found between the number of symptomatic infectious individuals (one of the crucial model responses) and three parameters which are effective contact rate with isolated infected individuals (*β*_*L*_), efficacy of face coverings (*ζ*) and face coverings compliance for our studied five countries. The public health implications of these findings are the dynamics of COVID-19 could be controlled by encouraging mass-level usage of efficacious face coverings. In addition, the high significance of *β*_*L*_ indicates that immediate isolation of detected patients is highly required.

**Figure 21:**
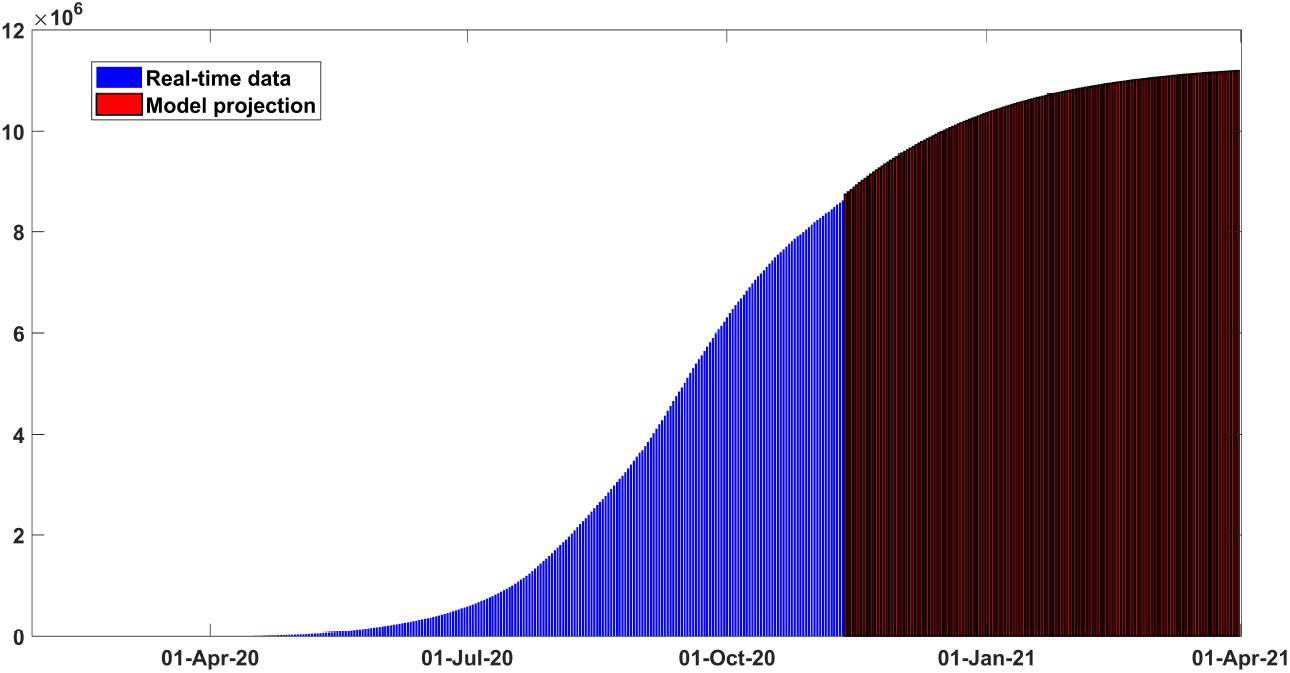
Projection results of cumulative cases for India from late January 2020 to late March 2021.

**Figure 22:**
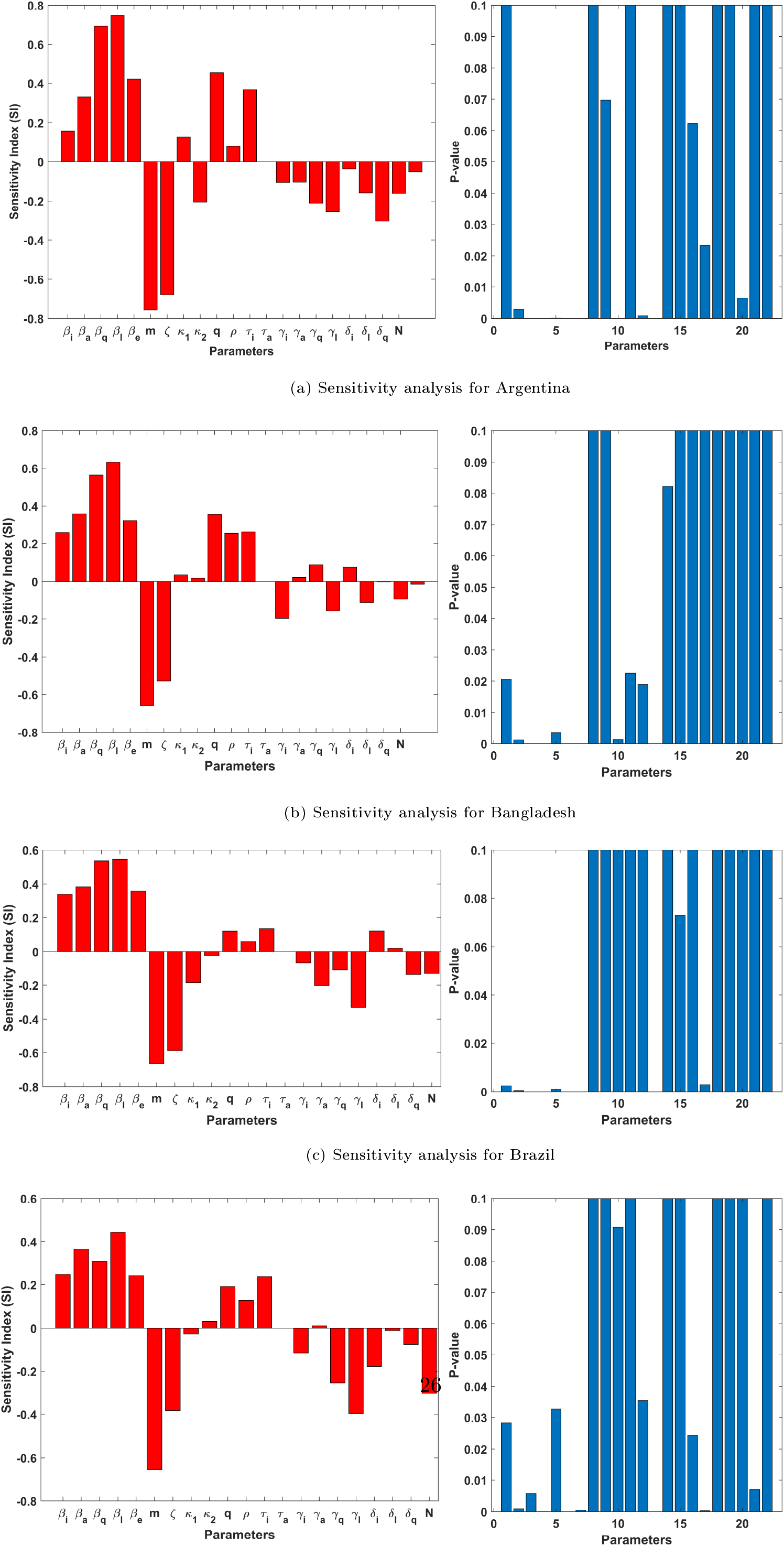

## 6. Solution of the model in Caputo-Fabrizio fractional derivative sense

### 6.1. Preliminaries

Here we recall the definitions of Caputo and Caputo-Fabrizio fractional derivatives.

#### Definition 1.

[32] The Caputo definition of non-integer order derivative of order *ϱ >* 0 of a function *G* : (0, *∞*) *→*ℝ is defined by

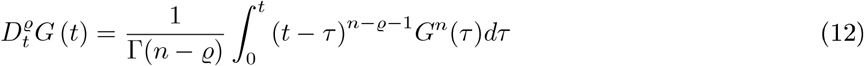

where *n* = [*ϱ*] + 1 and [*ϱ*] is the integer part of *ϱ*.

#### Definition 2.

[33, 34] For *G ∈ H*^1^(*c, d*) and 0 *< ϱ <* 1, the Caputo-Fabrizio (CF) fractional derivative (FD) of order *ϱ* is defined by

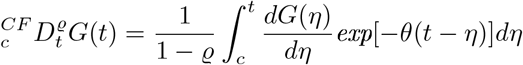

where 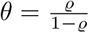

The CF non-integer order integral is defined as

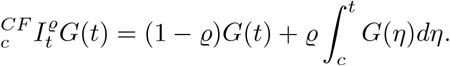

### 6.2. Existence and uniqueness analysis

Now, we prove the existence of unique solution for the given COVID-19 model in the sense of Caputo-Fabrizio fractional derivative by the application of fixed-point theory. In this concern, the proposed system can be rewritten in the equivalent form as follows:

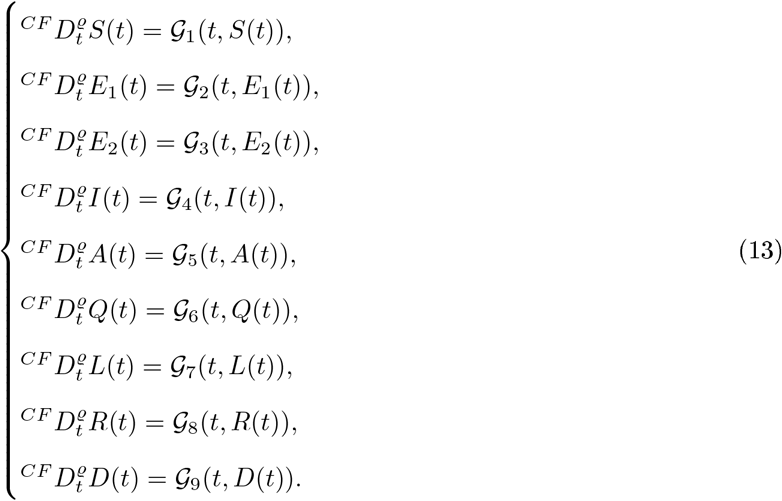

By applying the CF non-integer order integral operator, the above system (13), reduces to the following integral equation of Volterra type of order 0 *< ϱ <* 1.

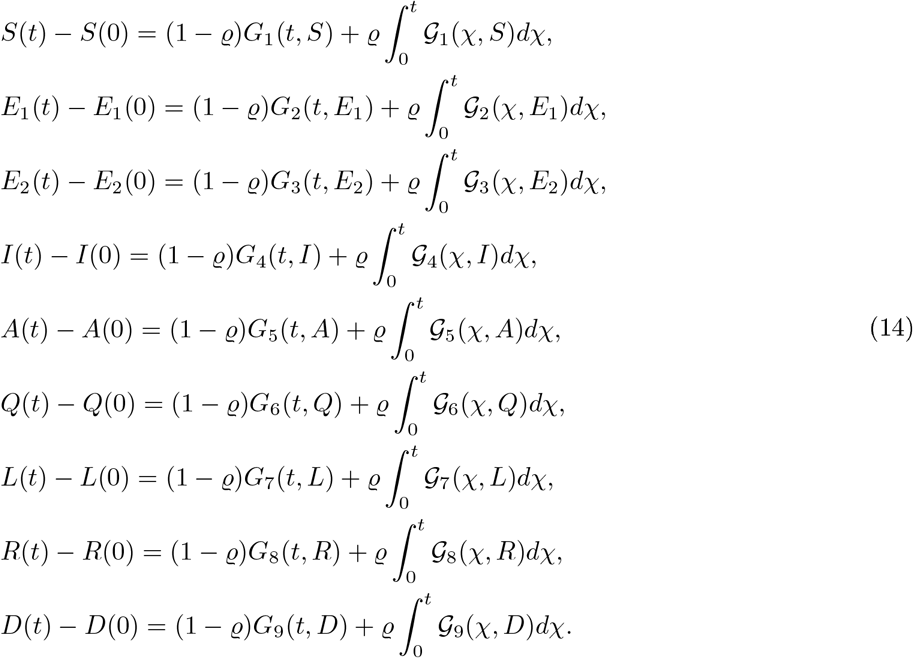

Now, we get the subsequent iterative algorithm

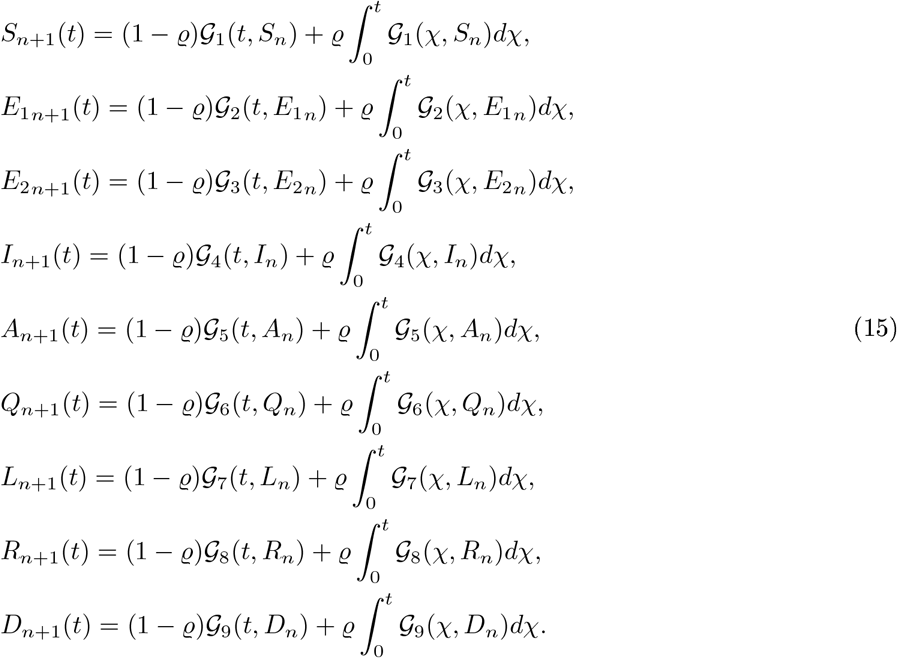

Here we assume that we can get the exact solution by taking the limit as n tends to infinity.

#### 6.2.1. Existence analysis by using Picard-Lindelof approach

Let us consider

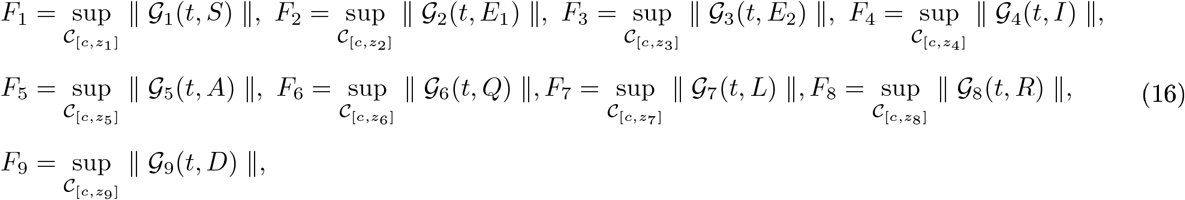

where

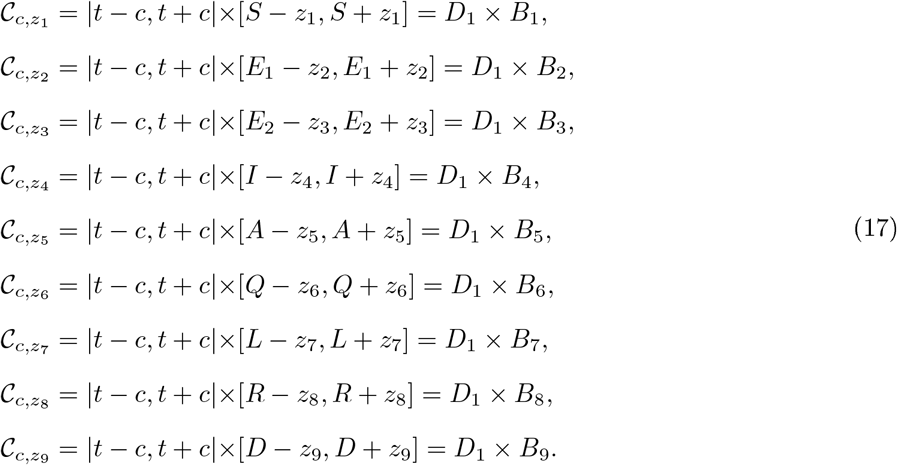

considering the Picard operator as

*ϕ* : *C*(*D*_1_, *B*_1_, *B*_2_, *B*_3_, *B*_4_, *B*_5_, *B*_6_, *B*_7_, *B*_8_, *B*_9_) *→ C*(*D*_1_, *B*_1_, *B*_2_, *B*_3_, *B*_4_, *B*_5_, *B*_6_, *B*_7_, *B*_8_, *B*_9_), given as follows:

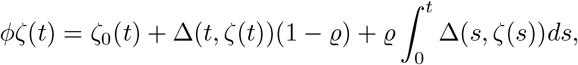

where 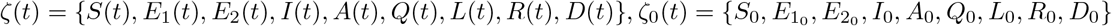 and Δ(*t, ζ*(*t*)) = *{𝒢*_1_(*t, S*(*t*)), *𝒢*_2_(*t, E*_1_(*t*)), *𝒢*_3_(*t, E*_2_(*t*)), *𝒢*_4_(*t, I*(*t*)), *𝒢*_5_(*t, A*(*t*)), *𝒢*_6_(*t, Q*(*t*)),

*𝒢*_7_(*t, L*(*t*)), *𝒢*_8_(*t, R*(*t*)), *𝒢*_9_(*t, D*(*t*))*}*. Next we assume that the solution of the non-integer order model are bounded within a time period, ‖ *ζ*(*t*) ‖_*∞*_ ≤ max*{z*_1_, *z*_2_, *z*_3_, *z*_4_, *z*_5_, *z*_6_, *z*_7_, *z*_8_, *z*_9_, *}*,

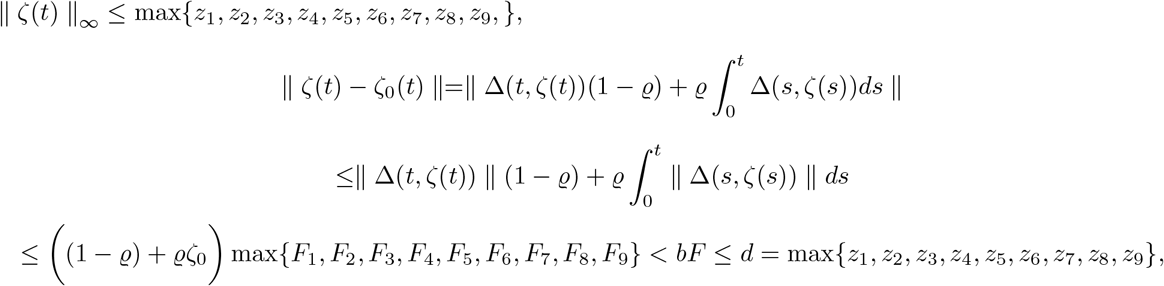

where we demand that

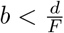. Now by the application of fixed point theorem pertaining to Banach space along with the metric, we obtain

‖ *ϕζ*_1_ *− ϕζ*_2_ ‖_*∞*_ = sup_*t→B*_ |*ζ*_1_ *− ζ*_2_|. Now we have

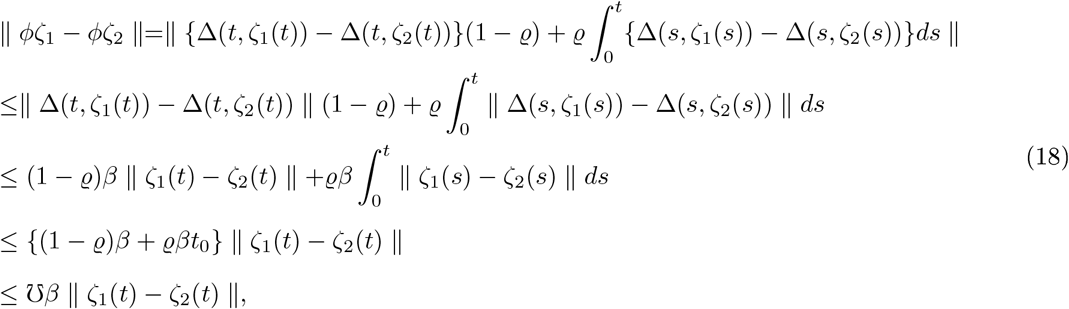

with *β <* 1. Since *ζ* is a contraction, we have *☋β <* 1, hence the given operator *ϕ* is also a contraction. Therefore, the model involving C-F derivative given in Eq. (13) has a unique set of solution.

### 6.3. Solution method in Caputo-Fabrizio Operator

We now derive the solution method for *I*(*t*) equation of the system (13) and for the rest of the equations it will be similar. The corresponding Volterra integral equation for *I*(*t*) is as follows.

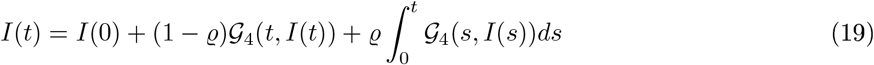

We have the following estimations at *t*_*k*_

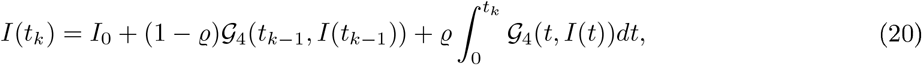

and at *t*_*k*+1_

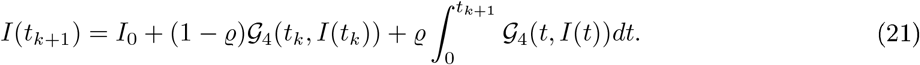

Subtracting equation (21) from (20), we obtain

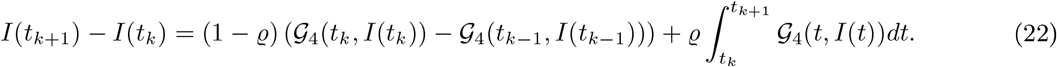

Then by linear interpolation about *𝒢*_4_(*t, I*(*t*)) and applying trapezoid rule for integration on the integral term, we can then write

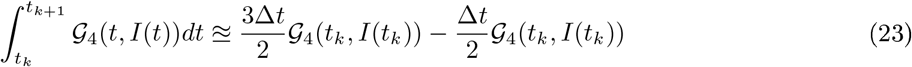

where Δ*t* = *t*_*k*_ *− t*_*k−*1_. Hence, we have the numerical approximation for equation of *I*(*t*) as

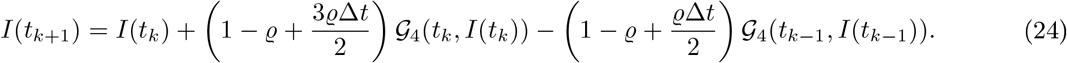

#### Theorem 5.

The numerical approximation (24) is unconditionally stable if

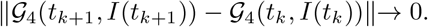

*Proof* Let *I*(*t*) be the solution of a differential equation as shown in (19) under CF non-integer order derivative operator sense. Then we have to evaluate the norm

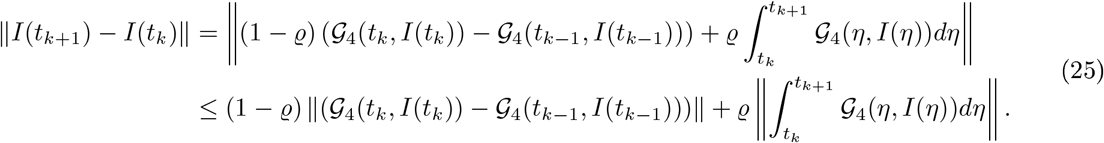

For *k → ∞*, we have

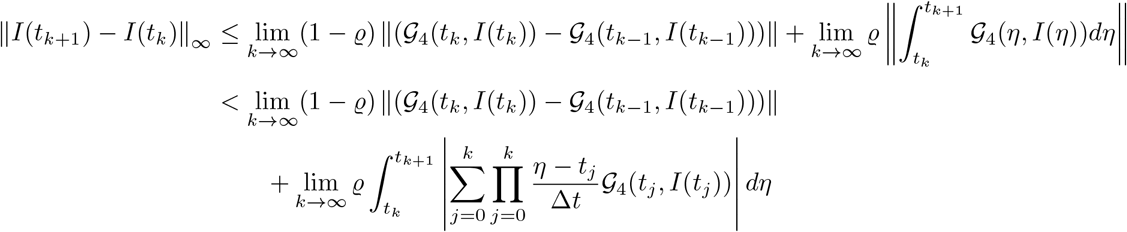

Clearly, the second term of the above inequality approaches zero when *k → ∞*. Now, if ||*𝒢*_4_(*t*_*k*+1_, *I*(*t*_*k*+1_))*− 𝒢*_4_(*t*_*k*_, *I*(*t*_*k*_))||*→* 0 as *k → ∞*, we educe that the numerical solution is stable.

### 6.4. Graphical simulations

After giving the all necessary theoretical concerns, now we do the graphical simulations via CF fractional derivative. In our paper, we are using the real numerical data of COVID-19 for five different countries named as Argentina, Bangladesh, Brazil, Colombia and India respectively. Here first we perform the graphs for Argentina COVID-19 cases. To perform numerical simulations, we use parameter values summarize in Table 2. In the family of Figure 23, we analysed the plots of *S*(*t*), *E*_1_(*t*), *E*_2_(*t*), *I*(*t*), *A*(*t*) and *Q*(*t*). We observed that for different fractional order values peaks are well defined and when we decrease the fractional order then the peaks sifted towards the later time period. In the group of Figs. 24-25, first we show the nature of *L*(*t*), *R*(*t*), *D*(*t*) and then analysed the plots of *I*(*t*) versus *S*(*t*), *A*(*t*), *Q*(*t*), *L*(*t*) and *R*(*t*). In the comparison of given classes with infected individuals, we see that when the infectious *I*(*t*) increases then asymptomatic infectious *A*(*t*) also increases with same nature. In sub-figs. 24d-25b, we see that the fractional order does not play any big role because the nature of the classes is nearly same at all different fractional order values *Q*. Initial values of given classes for Argentina are *S*(0) = 45333107, *E*_1_(0) = 10, *E*_2_(0) = 4, *I*(0) = 2, *A*(0) = 1, *Q*(0) = 0, *L*(0) = 0, *R*(0) = 0 and *D*(0) = 0. We have used the total population of the country for *S*(0).

**Figure 23:**
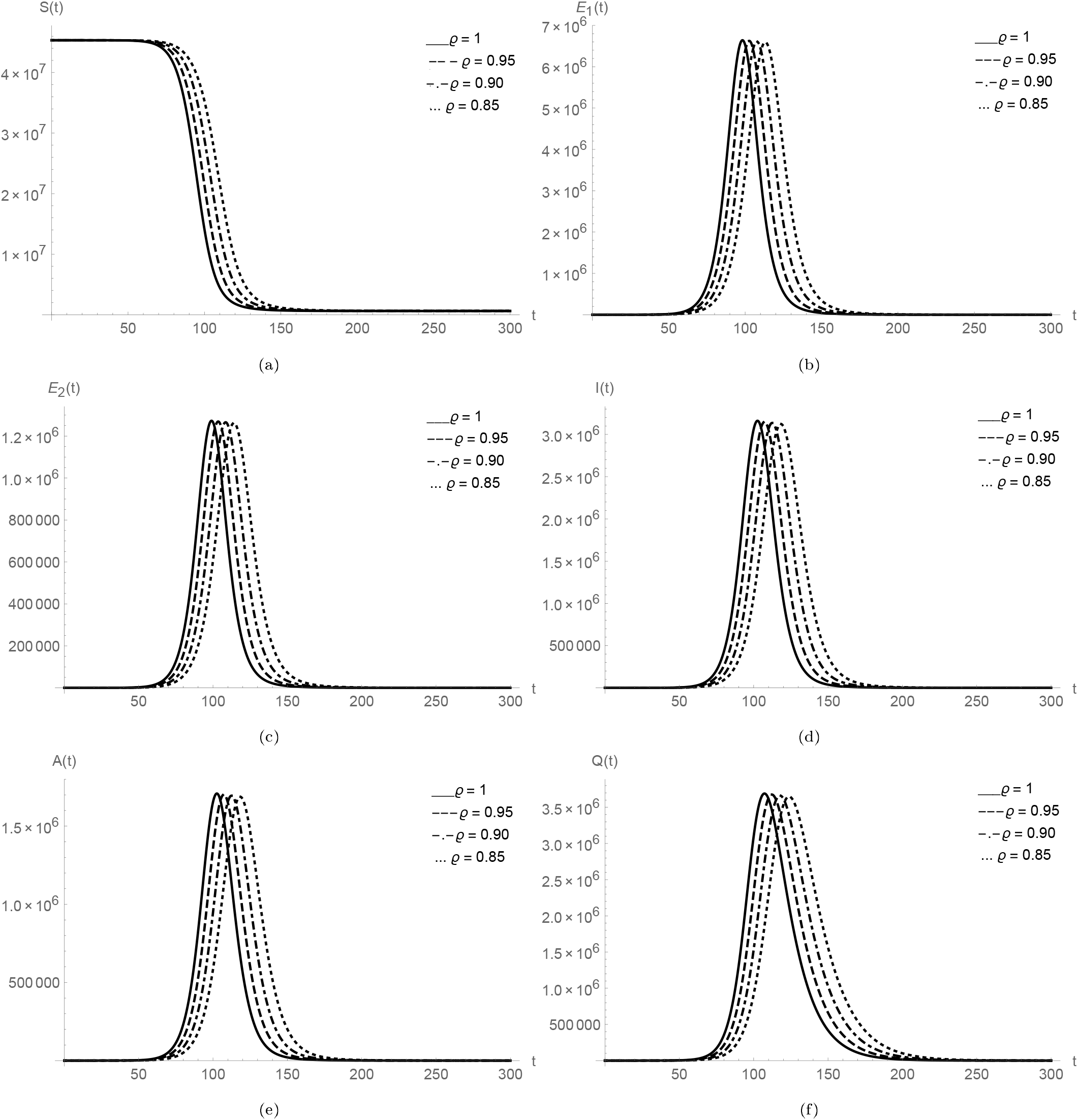
Plots of *S*(*t*), *E*_1_(*t*), *E*_2_(*t*), *I*(*t*), *A*(*t*) and *Q*(*t*) for Argentina data.

**Figure 24:**
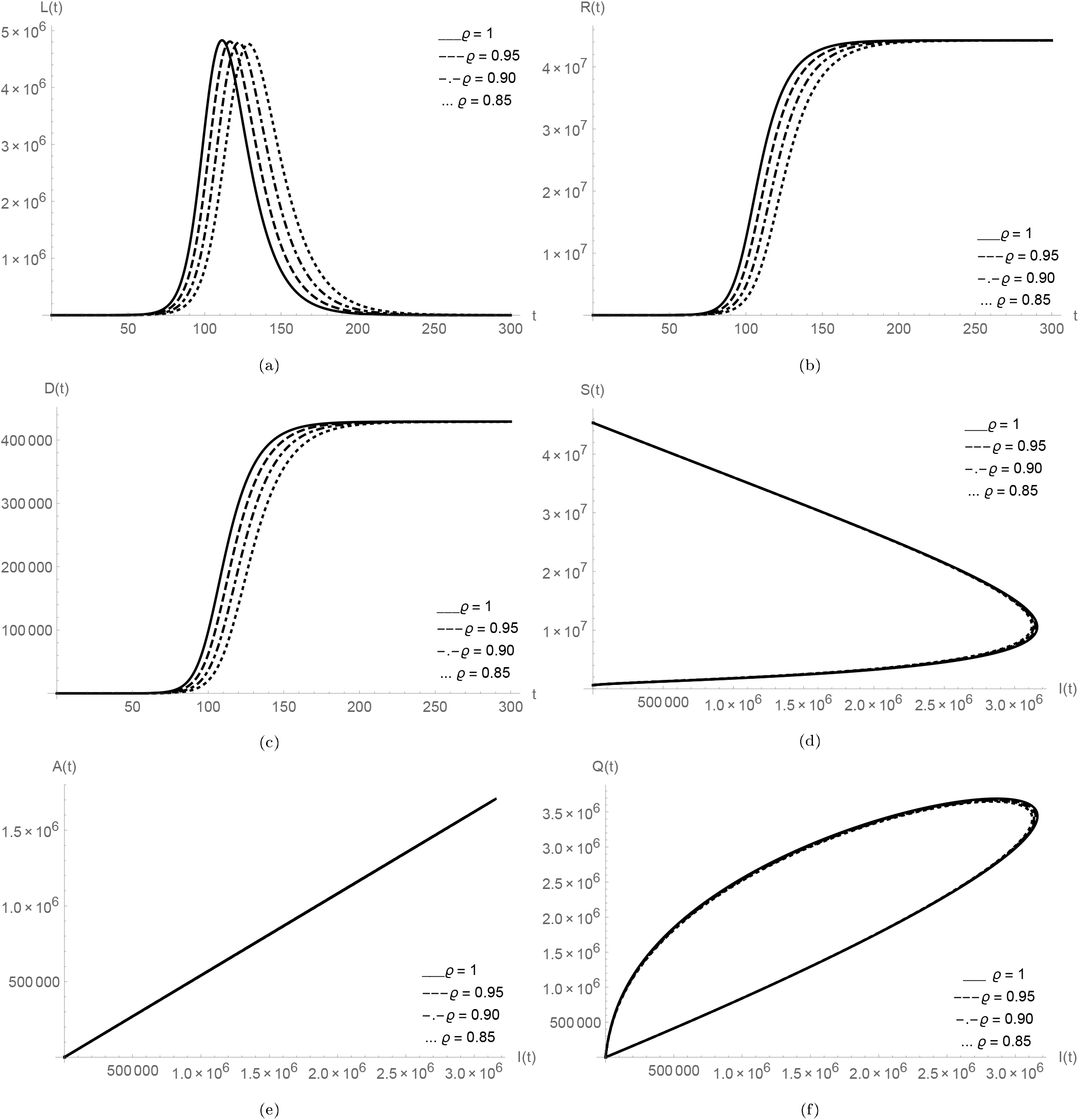
Plots of *L*(*t*), *R*(*t*), *D*(*t*) and relationship of *I*(*t*) versus *S*(*t*), *A*(*t*), *Q*(*t*), *L*(*t*), *R*(*t*) for Argentina data.

Now we perform the graphs for COVID-19 cases in Bangladesh. To perform numerical simulations, we use parameter values summarize in Table 3. In the family of Figure 26, we analysed the plots of *S*(*t*), *E*_1_(*t*), *E*_2_(*t*), *I*(*t*), *A*(*t*) and *Q*(*t*). We observed that for different fractional order values peaks are well defined and when we decrease the fractional order then the peaks sifted towards the later time period. In the group of Figures 27-28, first we show the nature of *L*(*t*), *R*(*t*), *D*(*t*) and then analysed the plots of *I*(*t*) versus *S*(*t*), *A*(*t*), *Q*(*t*), *L*(*t*) and *R*(*t*). In the comparison of given classes with infected individuals, we see that the nature of infectious *I*(*t*) is same as for Argentina, as when the population of infected individuals increases then asymptomatic infectious *A*(*t*) also increases with same nature. In sub-figures 27d-28b, we see that the fractional order does not play any big role because the nature of the classes is nearly same at all different fractional order values *ϱ*. Initial values of given classes for Bangladesh are *S*(0) = 164689383, *E*_1_(0) = 10, *E*_2_(0) = 4, *I*(0) = 2, *A*(0) = 1, *Q*(0) = 0, *L*(0) = 0, *R*(0) = 0 and *D*(0) = 0. We have used the total population of the country for *S*(0).

**Figure 25:**
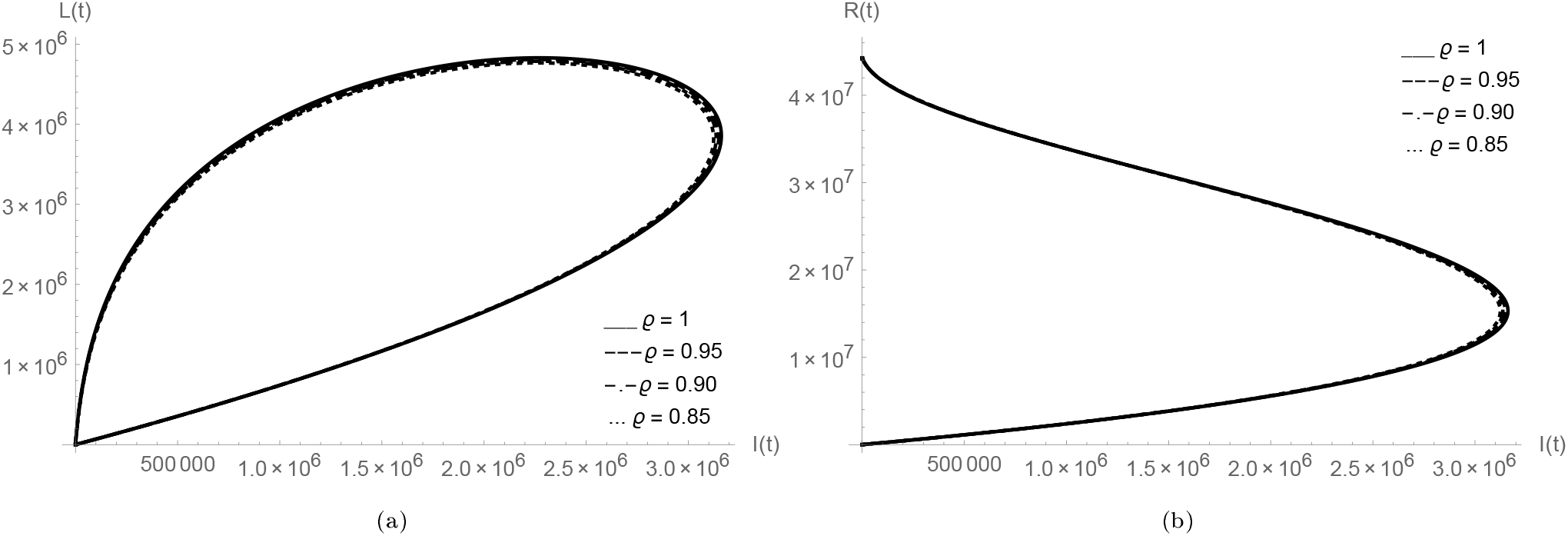
*I*(*t*) versus *L*(*t*), *R*(*t*) for Argentina data.

**Figure 26:**
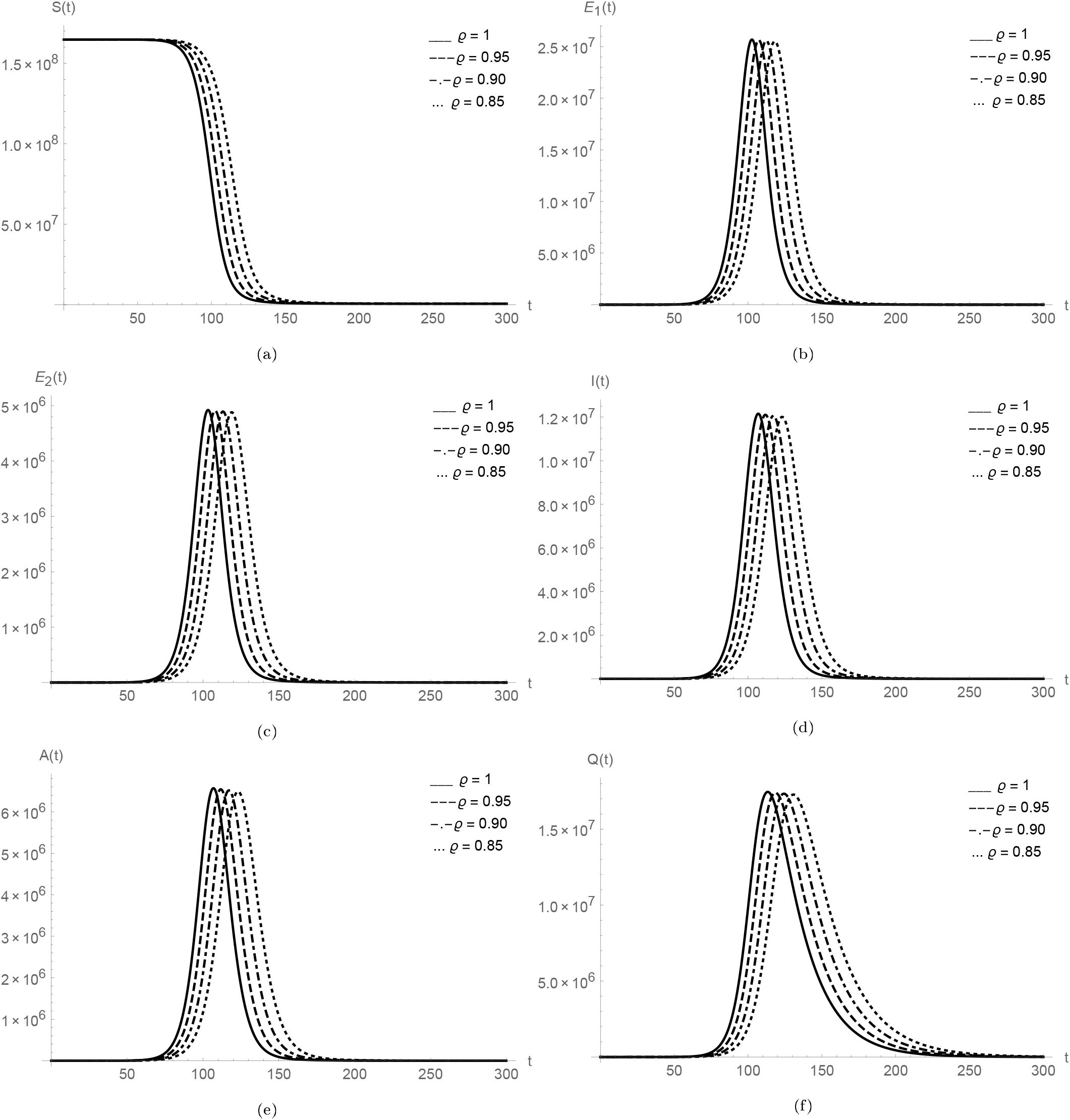
Plots of *S*(*t*), *E*_1_(*t*), *E*_2_(*t*), *I*(*t*), *A*(*t*) and *Q*(*t*) for Bangladesh data.

**Figure 27:**
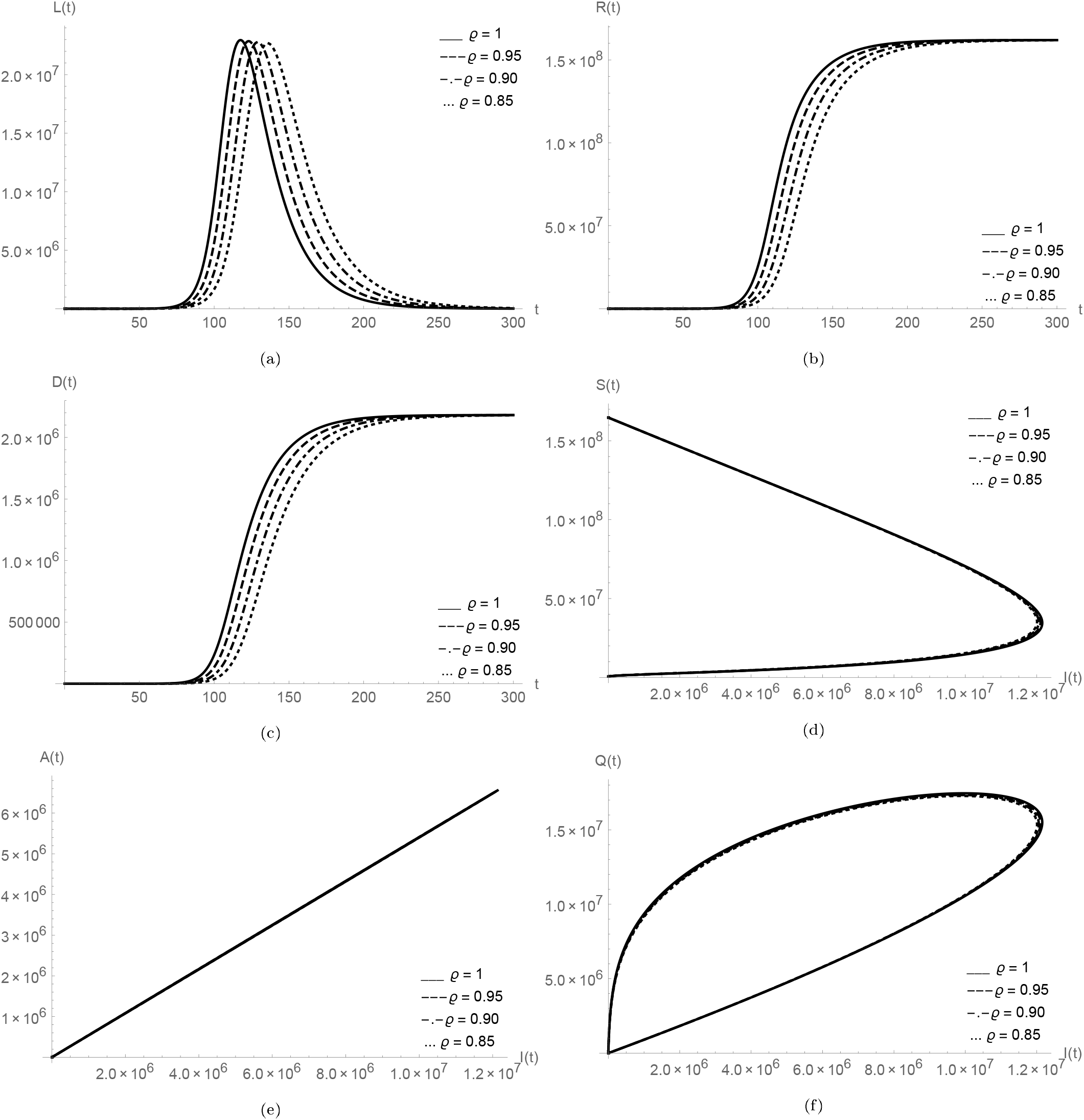
Plots of *L*(*t*);*R*(*t*);*D*(*t*) and relationship of *I*(*t*) versus *S*(*t*);*A*(*t*);*Q*(*t*);*L*(*t*);*R*(*t*) for Bangladesh data.

In the family of Figures 29-31, we exemplified the graphs for COVID-19 cases in Brazil. To perform numerical simulations, we use parameter values summarize in Table 4. In the family of Figure 29, we analysed the plots of *S*(*t*), *E*_1_(*t*), *E*_2_(*t*), *I*(*t*), *A*(*t*) and *Q*(*t*). We observed that for different fractional order values peaks are well defined and when we decrease the fractional order then the peaks sifted towards the later time period. In the collection of Figures 30-31, first we show the nature of *L*(*t*), *R*(*t*), *D*(*t*) and then analysed the plots of *I*(*t*) versus *S*(*t*), *A*(*t*), *Q*(*t*), *L*(*t*) and *R*(*t*). In the comparison of given classes with *I*(*t*), we again observed that the nature of infectious *I*(*t*) is same as for above countries, as when the population of infected individuals increases then asymptomatic infectious *A*(*t*) also increases with same nature. In sub-figures 30d-31b, we see that the fractional order does not play any big role because the nature of the classes is nearly same at all different fractional order values *ϱ*. Initial values of given classes for Brazil are *S*(0) = 212559417, *E*_1_(0) = 10, *E*_2_(0) = 4, *I*(0) = 2, *A*(0) = 1, *Q*(0) = 0, *L*(0) = 0, *R*(0) = 0 and *D*(0) = 0. We have used the total population of the country for *S*(0).

**Figure 28:**
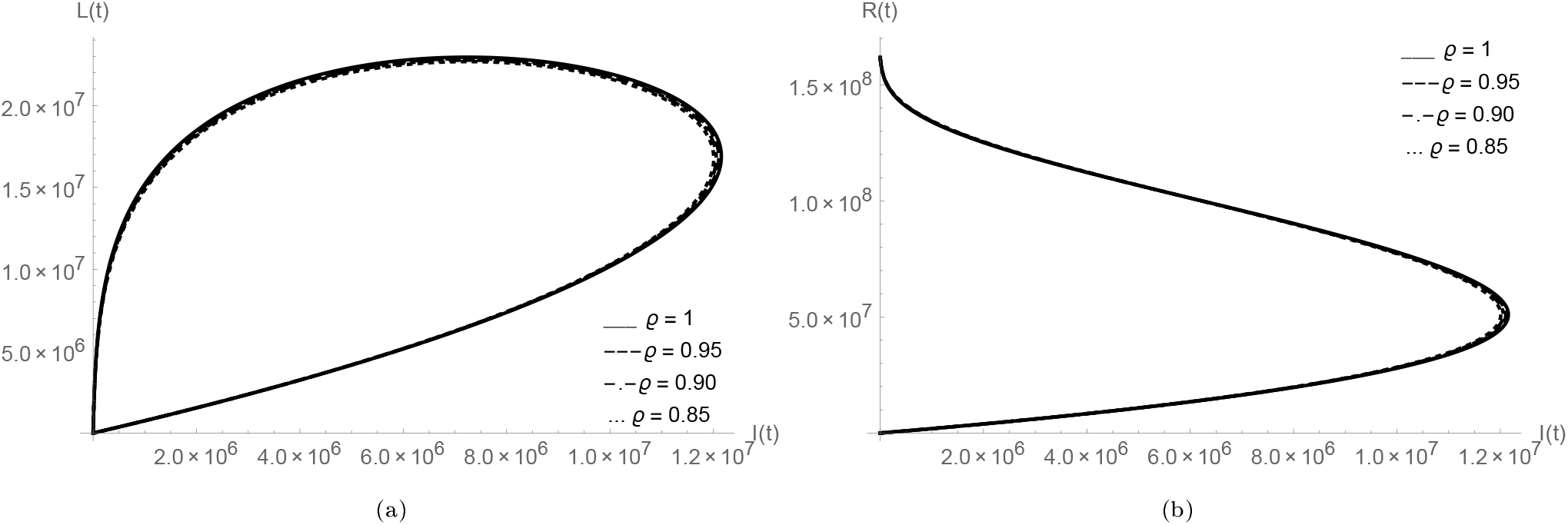
*I*(*t*) versus *L*(*t*), *R*(*t*) for Bangladesh data.

**Figure 29:**
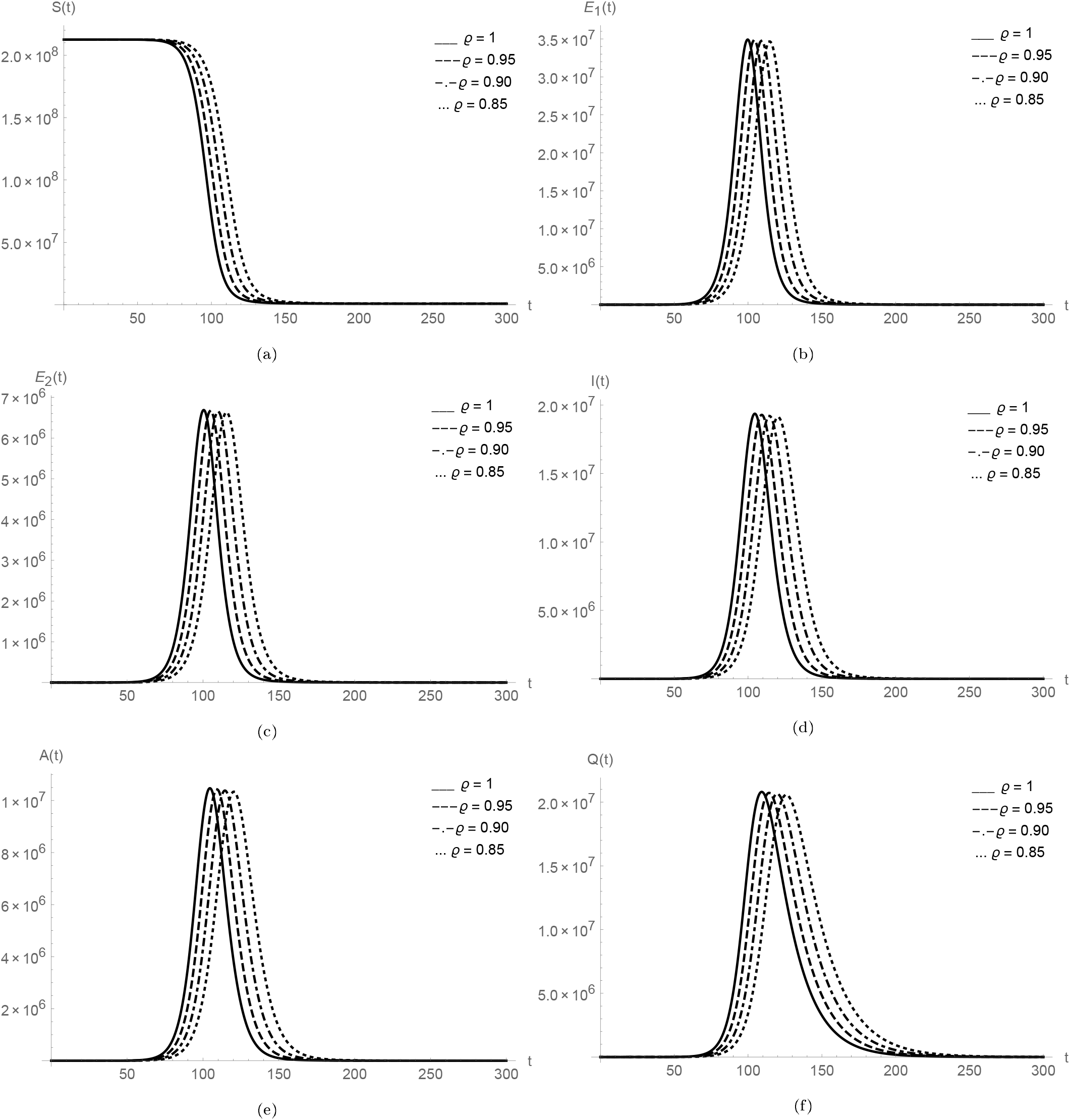
Plots of *S*(*t*), *E*_1_(*t*), *E*_2_(*t*), *I*(*t*), *A*(*t*) and *Q*(*t*) for Brazil data.

**Figure 30:**
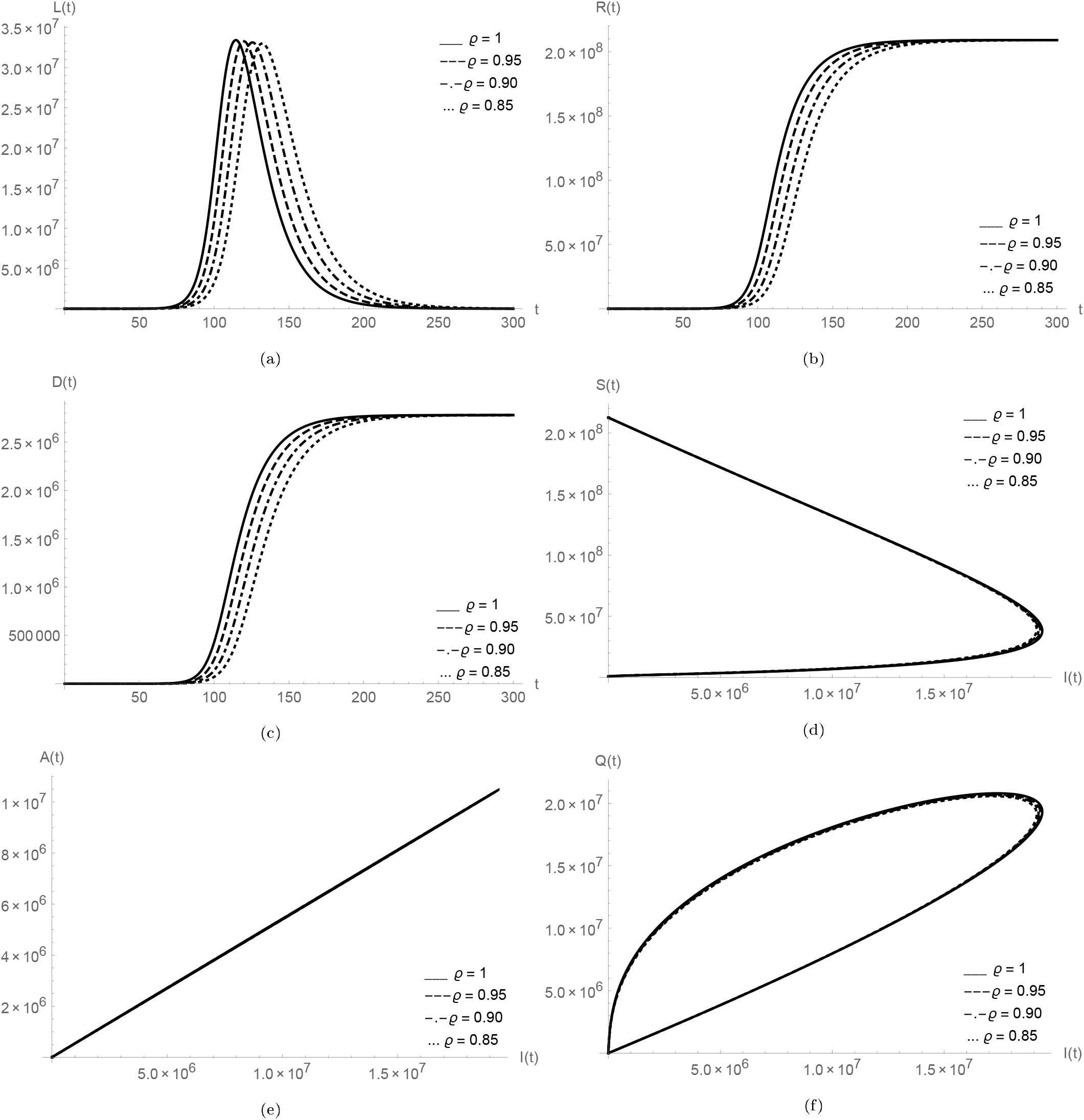
Plots of *L*(*t*), *R*(*t*), *D*(*t*) and relationship of *I*(*t*) versus *S*(*t*), *A*(*t*), *Q*(*t*), *L*(*t*), *R*(*t*) for Brazil data.

To continue the graphical simulations for the above mentioned countries to study the outbreaks of COVID-19, in the family of Figures 32-34, we exemplified the graphs for COVID-19 cases in Colombia. To perform numerical simulations, we have taken the numerical values summarize in Table 5. In the family of Figure 32, we analysed the plots of *S*(*t*), *E*_1_(*t*), *E*_2_(*t*), *I*(*t*), *A*(*t*) and *Q*(*t*). We observed that the nature of peaks is mostly same as for other above analysed data, for different fractional order values peaks are well defined and when we decrease the fractional order then the peaks sifted towards the later time period. In the collection of Figures 33-34, first we show the nature of *L*(*t*), *R*(*t*), *D*(*t*) and then analysed the plots of *I*(*t*) versus *S*(*t*), *A*(*t*), *Q*(*t*), *L*(*t*) and *R*(*t*). When we compare the given classes with *I*(*t*), we again observed that when the population of infected individuals increases then asymptomatic infectious *A*(*t*) also increases with same nature. In sub-figures 33d-34b, we see that the fractional order does not play any big role because the nature of the classes is nearly same at all different fractional order values *ϱ*. Initial values of given classes for Colombia are *S*(0) = 50882891, *E*_1_(0) = 10, *E*_2_(0) = 4, *I*(0) = 2, *A*(0) = 1, *Q*(0) = 0, *L*(0) = 0, *R*(0) = 0 and *D*(0) = 0. We have used the total population of the country for *S*(0).

**Table 5:**
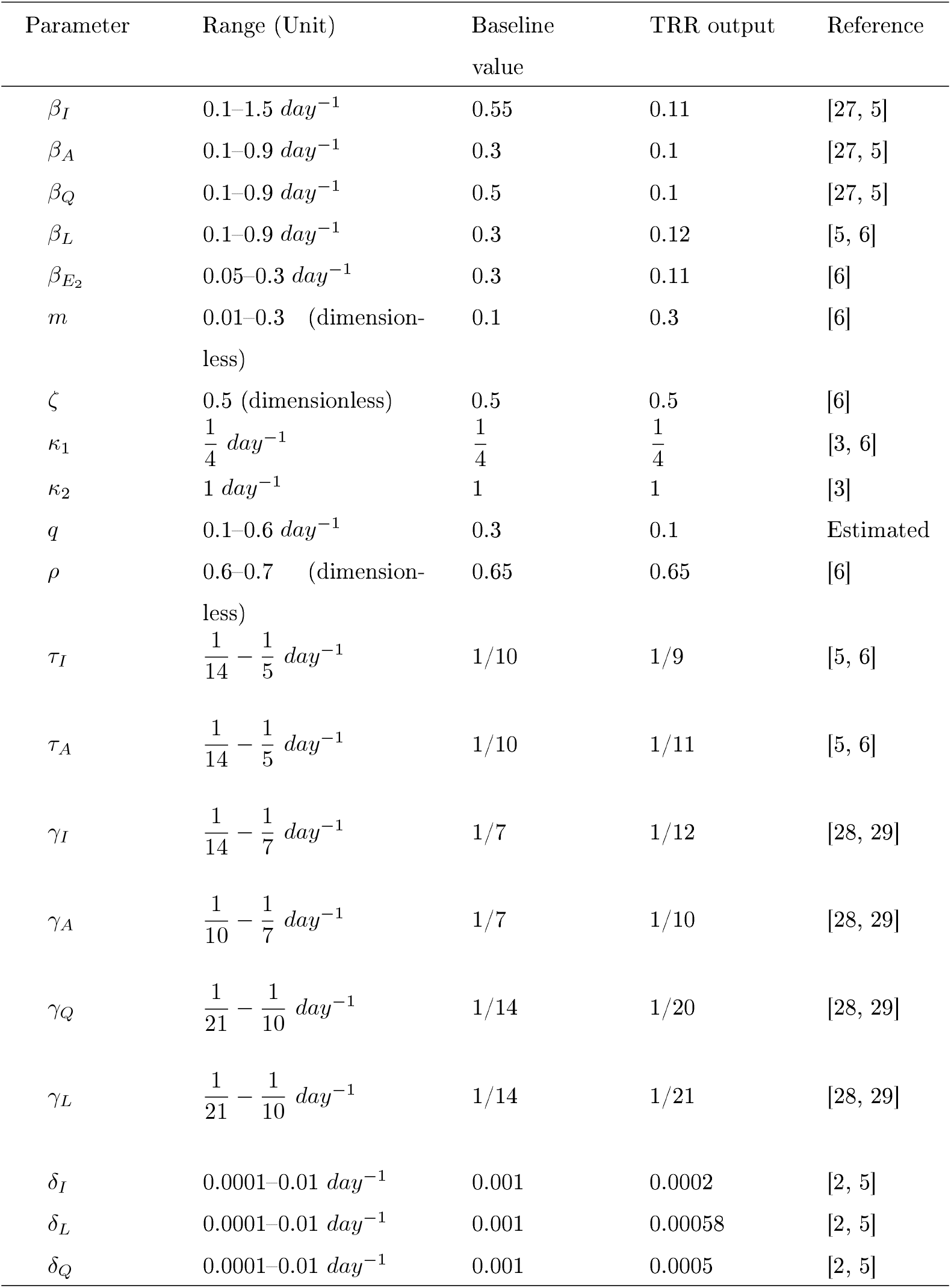
Calibrated parameters of the proposed model (1) using trust-region-re ective algorithm and daily COVID-19 cases data of Colombia

| Parameter | Range (Unit) | Baseline<br>value | TRR output | Reference |
| --- | --- | --- | --- | --- |
| $\beta_I$ | 0.1–1.5 $day^{-1}$ | 0.55 | 0.11 | [27, 5] |
| $\beta_A$ | 0.1–0.9 $day^{-1}$ | 0.3 | 0.1 | [27, 5] |
| $\beta_Q$ | 0.1–0.9 $day^{-1}$ | 0.5 | 0.1 | [27, 5] |
| $\beta_L$ | 0.1–0.9 $day^{-1}$ | 0.3 | 0.12 | [5, 6] |
| $\beta_{E_2}$ | 0.05–0.3 $day^{-1}$ | 0.3 | 0.11 | [6] |
| $m$ | 0.01–0.3 (dimensionless) | 0.1 | 0.3 | [6] |
| $\zeta$ | 0.5 (dimensionless) | 0.5 | 0.5 | [6] |
| $\kappa_1$ | $\frac{1}{4} day^{-1}$ | $\frac{1}{4}$ | $\frac{1}{4}$ | [3, 6] |
| $\kappa_2$ | 1 $day^{-1}$ | 1 | 1 | [3] |
| $q$ | 0.1–0.6 $day^{-1}$ | 0.3 | 0.1 | Estimated |
| $\rho$ | 0.6–0.7 (dimensionless) | 0.65 | 0.65 | [6] |
| $\tau_I$ | $\frac{1}{14} - \frac{1}{5} day^{-1}$ | 1/10 | 1/9 | [5, 6] |
| $\tau_A$ | $\frac{1}{14} - \frac{1}{5} day^{-1}$ | 1/10 | 1/11 | [5, 6] |
| $\gamma_I$ | $\frac{1}{14} - \frac{1}{7} day^{-1}$ | 1/7 | 1/12 | [28, 29] |
| $\gamma_A$ | $\frac{1}{10} - \frac{1}{7} day^{-1}$ | 1/7 | 1/10 | [28, 29] |
| $\gamma_Q$ | $\frac{1}{21} - \frac{1}{10} day^{-1}$ | 1/14 | 1/20 | [28, 29] |
| $\gamma_L$ | $\frac{1}{21} - \frac{1}{10} day^{-1}$ | 1/14 | 1/21 | [28, 29] |
| $\delta_I$ | 0.0001–0.01 $day^{-1}$ | 0.001 | 0.0002 | [2, 5] |
| $\delta_L$ | 0.0001–0.01 $day^{-1}$ | 0.001 | 0.00058 | [2, 5] |
| $\delta_Q$ | 0.0001–0.01 $day^{-1}$ | 0.001 | 0.0005 | [2, 5] |

**Figure 31:**
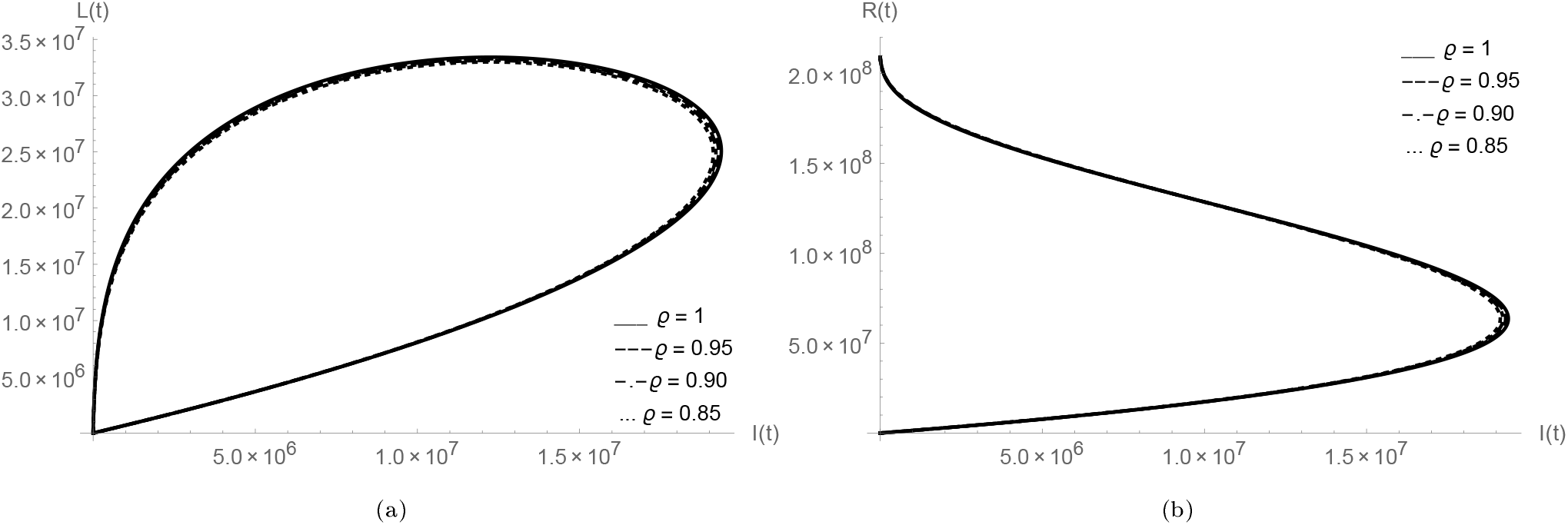
*I*(*t*) versus *L*(*t*), *R*(*t*) for Brazil data.

**Figure 32:**
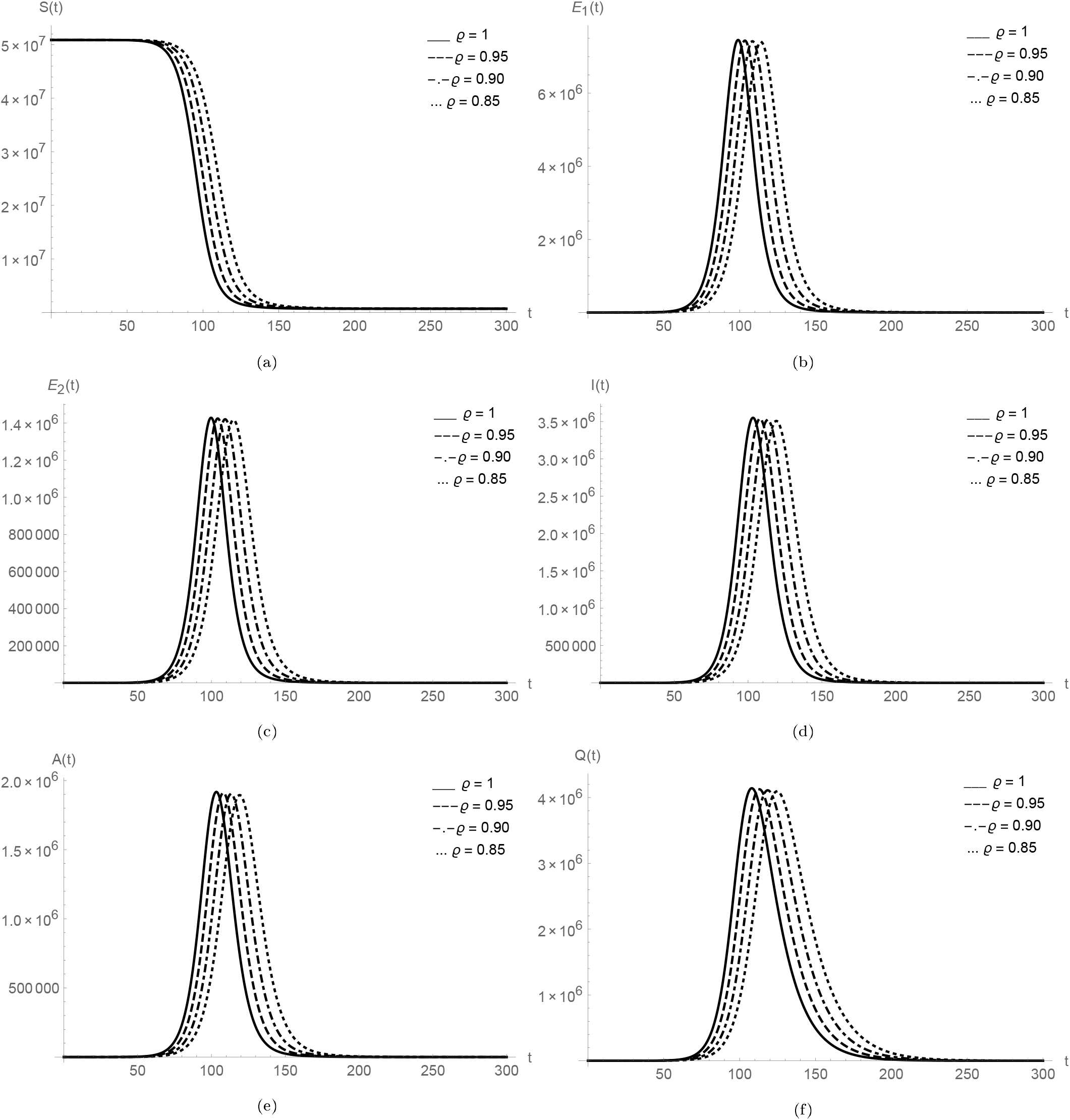
Plots of *S*(*t*), *E*_1_(*t*), *E*_2_(*t*), *I*(*t*), *A*(*t*) and *Q*(*t*) for Colombia data.

**Figure 33:**
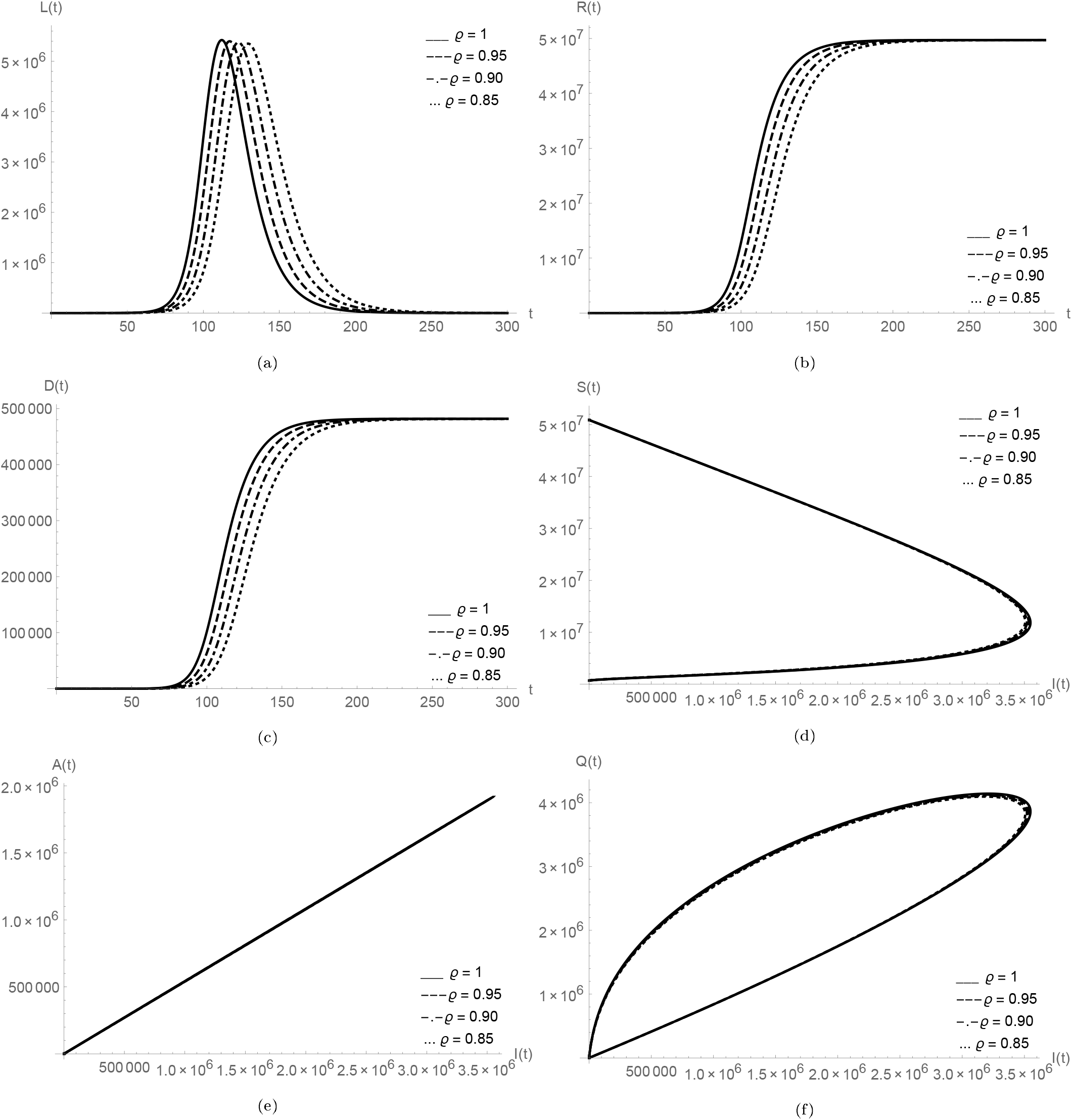
Plots of *L*(*t*), *R*(*t*), *D*(*t*) and relationship of *I*(*t*) versus *S*(*t*), *A*(*t*), *Q*(*t*), *L*(*t*), *R*(*t*) for Colombia data.

Eventually, we have done the graphical simulations for India which is the second highest populous country and also the second worst-hit country by COVID-19. To study the outbreak of COVID-19 in India, in the family of Figures 35-37, we exemplified the all necessary graphs of given classes to observe the dynamics of COVID-19. To perform numerical simulations, we took the numerical values from the Table 6. In the family of Figure 35, we analysed the plots of *S*(*t*), *E*_1_(*t*), *E*_2_(*t*), *I*(*t*), *A*(*t*) and *Q*(*t*). We observed that the nature of peaks is mostly same as for other above analysed data, for different fractional order values peaks are well defined and when we decrease the fractional order then the peaks sifted towards the later time period. In the collection of Figures 36-37, first we show the nature of *L*(*t*), *R*(*t*), *D*(*t*) and then analysed the plots of *I*(*t*) versus *S*(*t*), *A*(*t*), *Q*(*t*), *L*(*t*) and *R*(*t*). When we compare the given classes with *I*(*t*), we again observed that when the population of infected individuals increases then asymptomatic infectious *A*(*t*) also increases with same nature. In sub-figures 36d-37b, we observed that at the different fractional order values the nature of the classes is nearly same. Initial values of given classes for India are *S*(0) = 414001316, *E*_1_(0) = 10, *E*_2_(0) = 4, *I*(0) = 2, *A*(0) = 1, *Q*(0) = 0, *L*(0) = 0, *R*(0) = 0 and *D*(0) = 0. We have used the 30% of the total population of India for *S*(0).

**Table 6:** Calibrated parameters of the proposed model (1) using trust-region-re ective algorithm and daily COVID-19 cases data of India

| Parameter | Range (Unit) | Baseline<br>value | TRR output | Reference |
| --- | --- | --- | --- | --- |
| $\beta_I$ | 0.1–1.5 $day^{-1}$ | 0.55 | 0.18 | [27, 5] |
| $\beta_A$ | 0.1–0.9 $day^{-1}$ | 0.3 | 0.13 | [27, 5] |
| $\beta_Q$ | 0.1–0.9 $day^{-1}$ | 0.5 | 0.2 | [27, 5] |
| $\beta_L$ | 0.1–0.9 $day^{-1}$ | 0.3 | 0.22 | [5, 6] |
| $\beta_{E_2}$ | 0.05–0.3 $day^{-1}$ | 0.3 | 0.15 | [6] |
| $m$ | 0.01–0.3 (dimension-<br>less) | 0.1 | 0.23 | [6] |
| $\zeta$ | 0.5 (dimensionless) | 0.5 | 0.5 | [6] |
| $\kappa_1$ | $\frac{1}{4} day^{-1}$ | $\frac{1}{4}$ | $\frac{1}{4}$ | [3, 6] |
| $\kappa_2$ | 1 $day^{-1}$ | 1 | 1 | [3] |
| $q$ | 0.1–0.6 $day^{-1}$ | 0.3 | 0.25 | Estimated |
| $\rho$ | 0.6–0.7 (dimension-<br>less) | 0.65 | 0.65 | [6] |
| $\tau_I$ | $\frac{1}{14} - \frac{1}{5} day^{-1}$ | 1/10 | 1/8 | [5, 6] |
| $\tau_A$ | $\frac{1}{14} - \frac{1}{5} day^{-1}$ | 1/10 | 1/9 | [5, 6] |
| $\gamma_I$ | $\frac{1}{14} - \frac{1}{7} day^{-1}$ | 1/7 | 1/12 | [28, 29] |
| $\gamma_A$ | $\frac{1}{10} - \frac{1}{7} day^{-1}$ | 1/7 | 1/10 | [28, 29] |
| $\gamma_Q$ | $\frac{1}{21} - \frac{1}{10} day^{-1}$ | 1/14 | 1/16 | [28, 29] |
| $\gamma_L$ | $\frac{1}{21} - \frac{1}{10} day^{-1}$ | 1/14 | 1/18 | [28, 29] |
| $\delta_I$ | 0.0001–0.01 $day^{-1}$ | 0.001 | 0.0002 | [2, 5] |
| $\delta_L$ | 0.0001–0.01 $day^{-1}$ | 0.001 | 0.00032 | [2, 5] |
| $\delta_Q$ | 0.0001–0.01 $day^{-1}$ | 0.001 | 0.0003 | [2, 5] |

**Figure 34:**
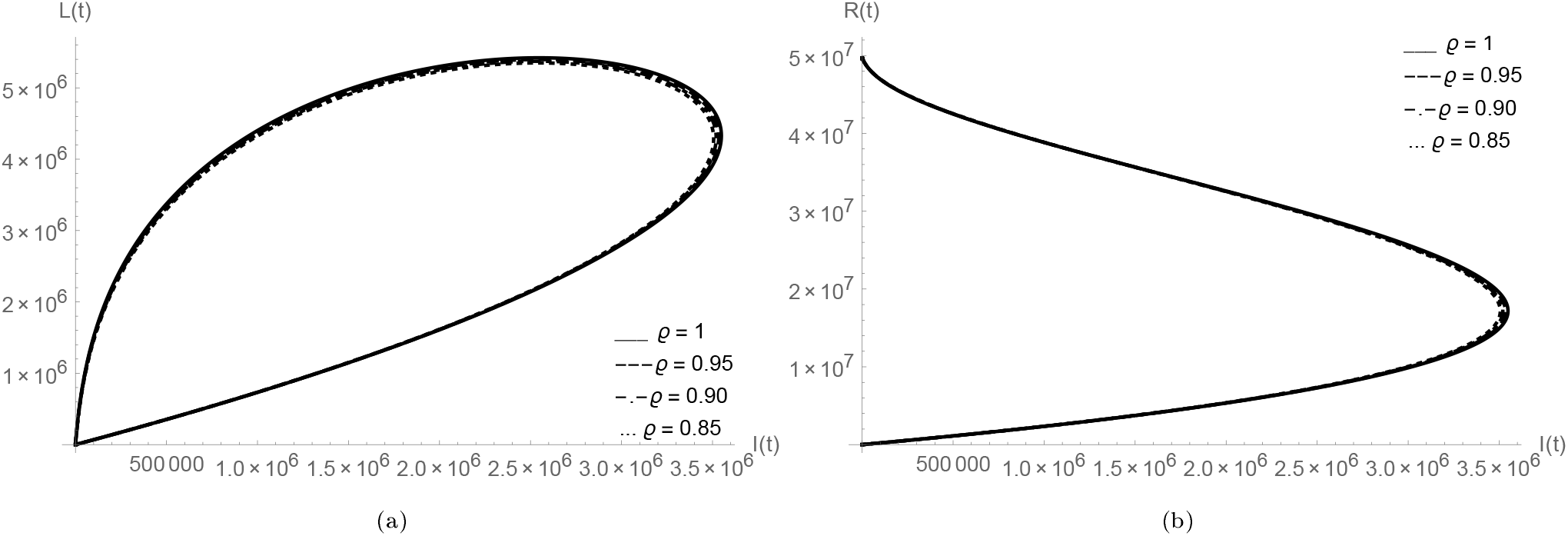
*I*(*t*) versus *L*(*t*), *R*(*t*) for Colombia data.

**Figure 35:**
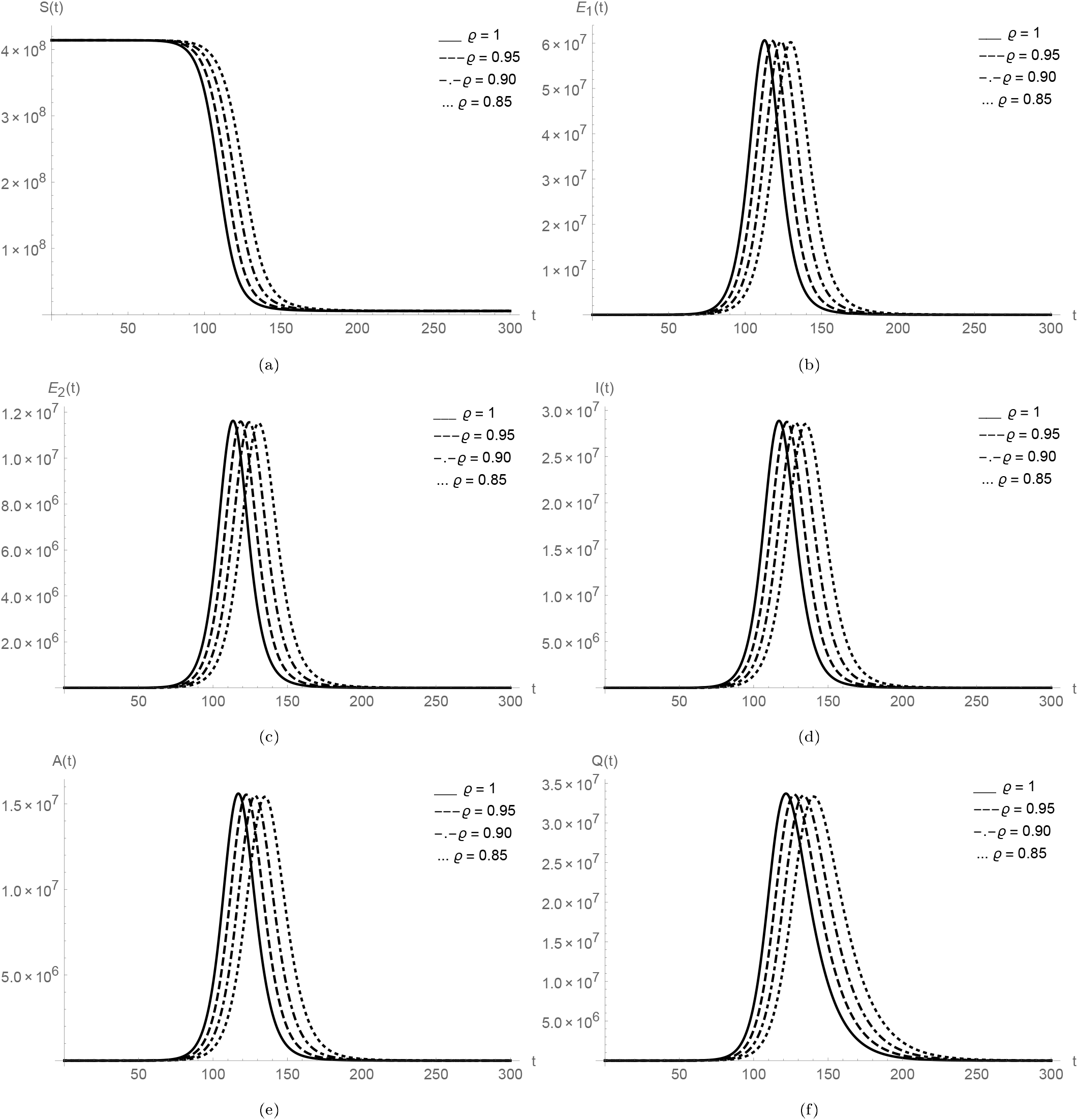
Plots of *S*(*t*), *E*_1_(*t*), *E*_2_(*t*), *I*(*t*), *A*(*t*) and *Q*(*t*) for India data.

**Figure 36:**
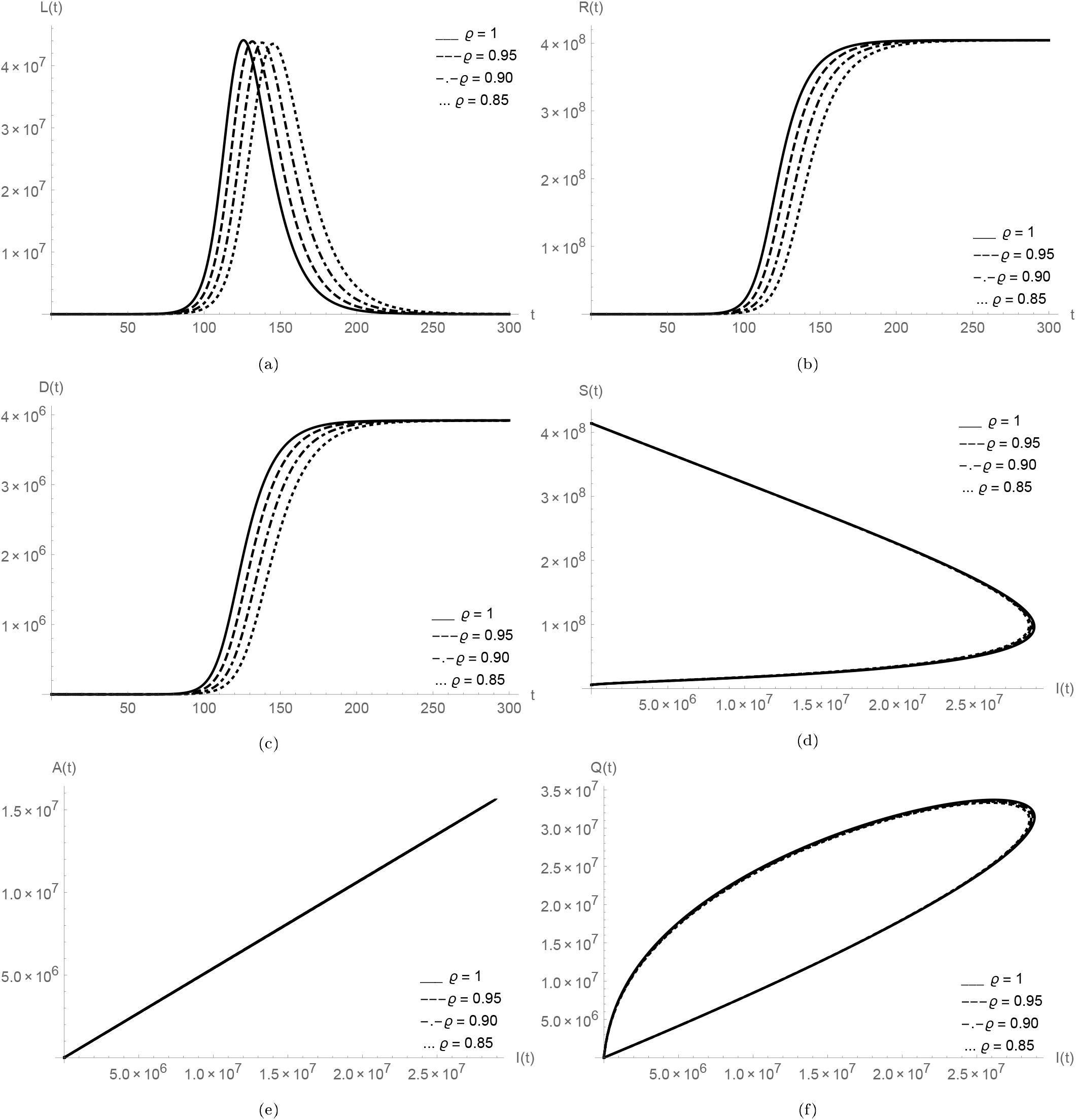
Plots of *L*(*t*), *R*(*t*), *D*(*t*) and relationship of *I*(*t*) versus *S*(*t*), *A*(*t*), *Q*(*t*), *L*(*t*), *R*(*t*) for India data.

**Figure 37:**
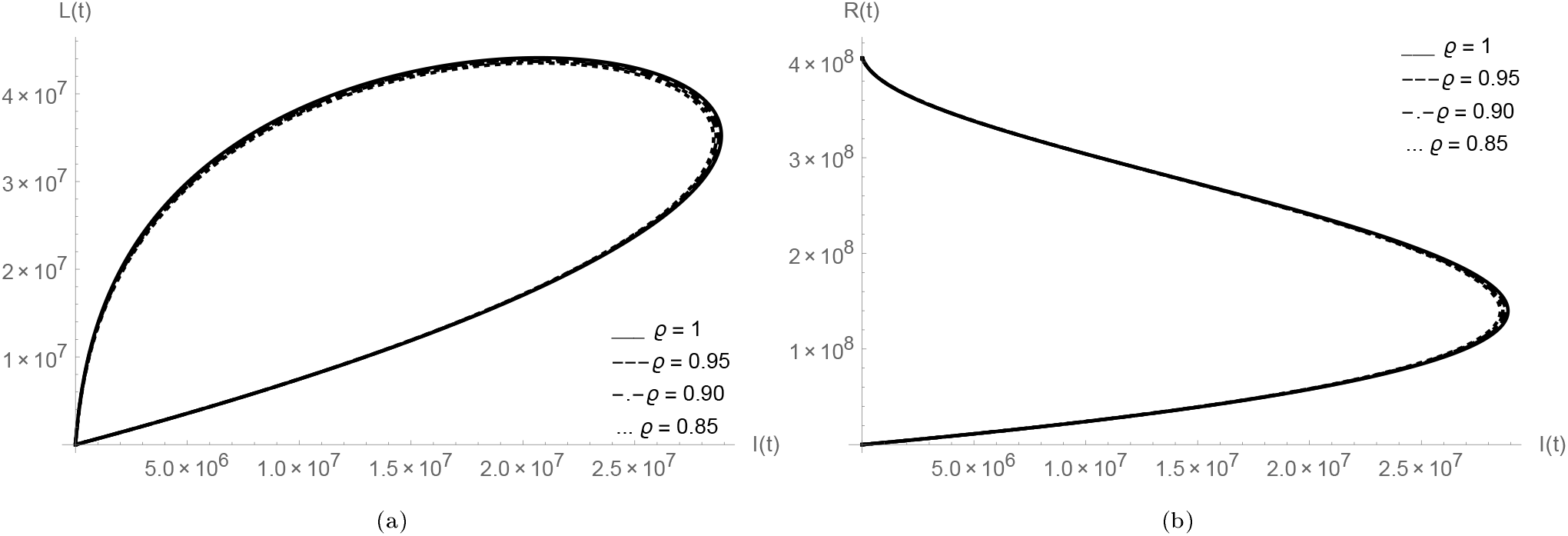
*I*(*t*) versus *L*(*t*), *R*(*t*) for India data.

From the all above graphical observations we found that the Caputo-Fabrizio fractional derivative playing well to study the outbreaks of coronavirus in the aforesaid five countries.

## 7. Optimal control problem formulation

In this concern, our main aim is to decrease the number of infected individuals with COVID-19 at the same time decrease the cost *J* (*v*) associated with their strategies. For this purpose, we use a control function *v* = (*v*_1_, *v*_2_, *v*_3_), where *v*_1_(*t*) is for introducing the public education or aware the public with health-care measures, *v*_2_(*t*) is the control function for enhancement of the strength of treatment for the infected individuals, *v*_3_(*t*) is the control function for the necessary suggestions of health care measures for those who are in asymptomatic infectious class and yet not admitted in the hospital.

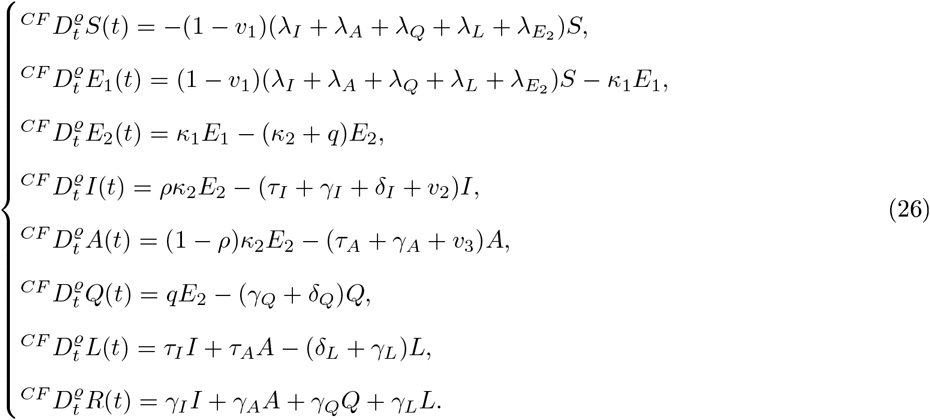

To define the optimal control problem (OCP), we are excluding the death equation *D*(*t*), because there is no significance of deaths in optimal controls.Now consider the state system given in (26) in *R*^8^, with the set of admissible control function. Ω = *{*(*v*_1_(.), *v*_2_(.), *v*_3_(.)|*v*_*i*_ *is Lebsegue measurable on*[0, 1] 0 ≤ (*v*_1_(.), *v*_2_(.), *v*_3_(.) ≤ 1 So the objective functional is defined by

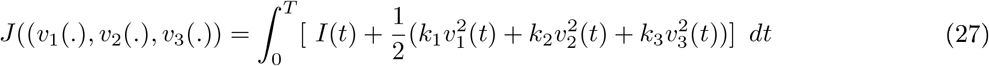

where the constants *k*_1_, *k*_2_ and *k*_3_ are a measure of associative cost with the controls *v*_1_, *v*_2_ and *v*_3_. Then we find the optimal controls *v*_1_, *v*_2_ and *v*_3_ to minimize the cost function.

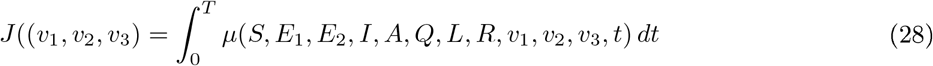

subject to constraint,

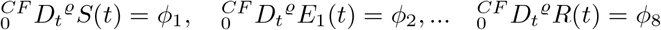

where *ϕ*_*j*_ = *ϕ*(*S, E*_1_, *E*_2_, *I, A, Q, L, R, V*_1_, *V*_2_, *V*_3_, *t*),*j* = 1, 2, …8 and the given initial coordination are agreed *S*(0) = *S*_0_, *E*_1_(0) = *E*_1_0, *R*(0) = *R*_0_

Now let us take the following modified cost function

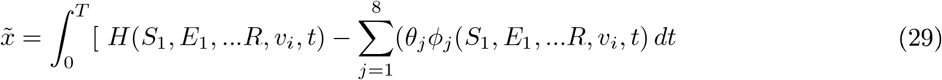

where *i* = 1, 2, 3 and *j* = 1, 2, 3…8 Hence the Hamiltonian is defined as follows:

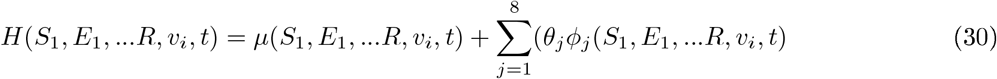

where *i* = 1, 2, 3 and *j* = 1, 2, 3…8 from Equation 29 and 30, the necessary and sufficient conditions for the functional optimal control problem (FOCP) are given as:

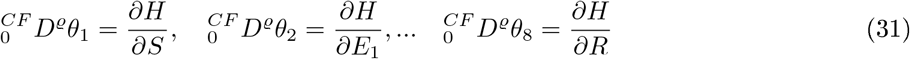

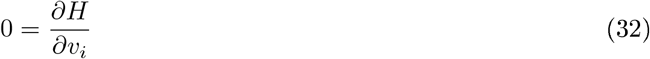

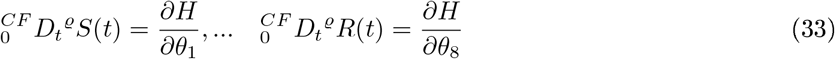

Moreover, *θ*_*j*_(*T*) = 0, *j* = 1, 2, …8, are the lagseuges multipliers Eqn 31 and 32 express the necessary condition in terms of a Hamiltonian for the OCP defined above.

### 7.1. Optimality conditions for fraction order

Let us write the Hamiltonian function as follows:

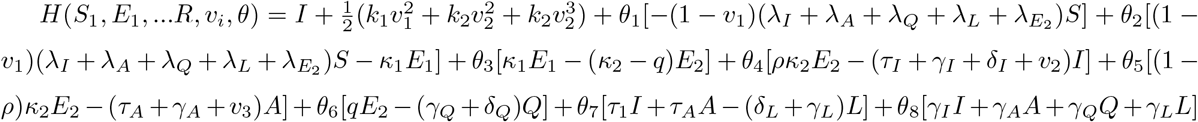

where *θ*_*j*_, *j* = 1, 2, …8, representing the lagragars multipliers called co-states.

#### Theorem

If 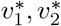 and 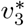 are optimal controls of the given OCP if 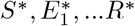 are corresponding optimal paths, then there exists co-state variables 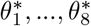, such that besides the given control system is satisfied, the following conditions are satisfied:

Co-state equations:

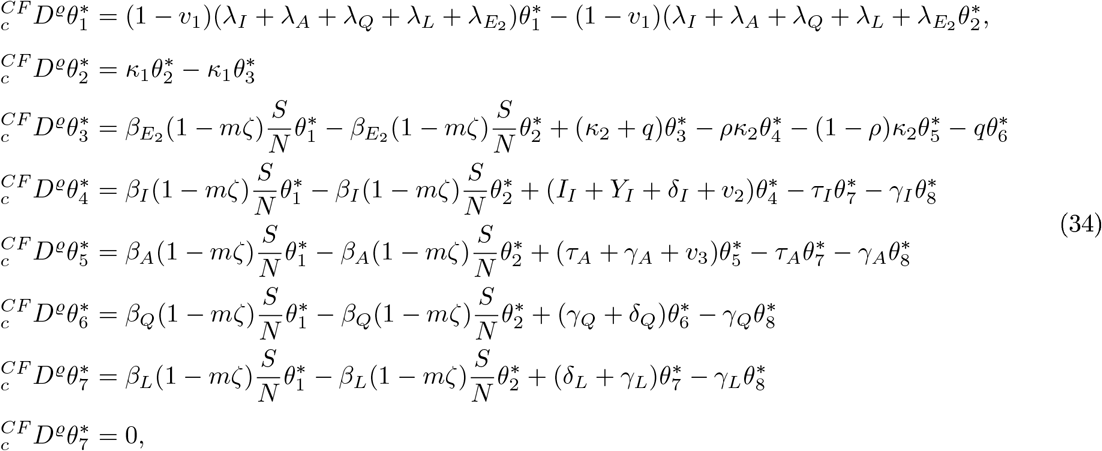

with transversality conditions *θ*_*j*_(*T*) = 0, *j* = 1, 2, …8 and optimality conditions given by

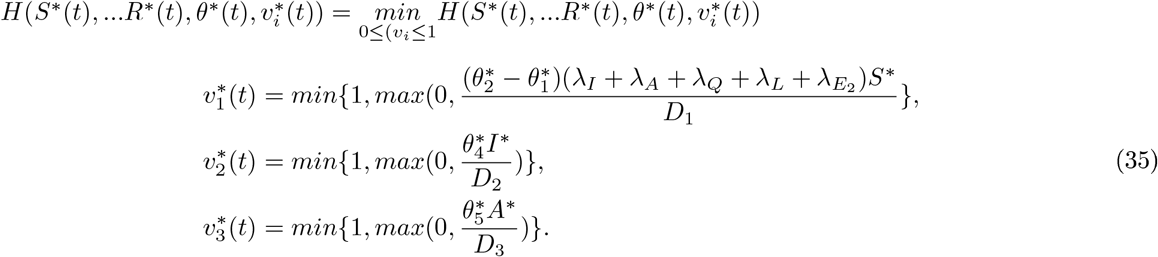

**Proof** The adjoint system (34) 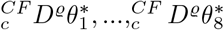 are obtained from the Hamiltonian *H* as

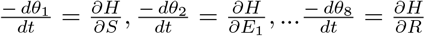

with zero final time conditions or transversality conditions,

*θ*_1_(*T*) = 0, *θ*_2_(*T*) = 0, …. and *θ*_8_(*T*) = 0 and the characteristic of the fractional optimal control given by (35) is obtained by solving the Eqn 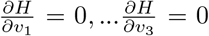 on the interior of the control set and using the property of control space *v*.

## 8. Conclusions

Different mathematical paradigms can provide considerable insights and scientific evidences pertinent to any ongoing epidemic dynamics. Based on those valuable information, health officials and public health experts can set up potential control strategies to battle against any epidemic. From the emergence of the novel coronavirus in China, researchers and scientists are working relentlessly to develop various mathematical modeling approaches to gain a deeper understanding on the progression dynamics of COVID-19 in the world. In addition, in the absence of any safe, effective and widely available COVID-19 vaccine, different preventive measures are the most effective tool in combating against the virulent virus. On the basis of robust forecasting results of reliable epidemiological models, government officials can deploy different public health intervention strategies to control the rapid transmission of the virus. In this chapter, a compartmental mathematical has been designed to describe the transmission dynamics of the COVID-19 incorporating all possible real-life interactions and effective non-pharmaceutical interventions. Disease-free equilibrium (DFE) of the proposed model is found to be globally asymptotically stable (GAS), whenever control reproduction number (*ℛ*_*c*_) less than unity. In addition, advanced forecasting techniques have also been applied for Argentina, Bangladesh, Brazil, Colombia and India to portray the future dynamics of the pandemic in near term. It has been enlightened in our study that mass-level using of highly effective face coverings could be a crucial factor in controlling the spread of coronavirus. Moreover, strict social-distancing measures and comprehensive contact-tracing are also effective strategies in battling against this pandemic. The public health implication of these insightful findings is government officials can undertake crucial clinical and public health decisions by analyzing all mathematical results and scientific evidences. Caputo-Fabrizio non-integer order derivative has been applied to solve the proposed mathematical model in fractional sense. We proved the existence of unique solution for the proposed fractional initial value problem. We proved the unconditional stability of the given technique. An important concern of fractional optimal control problem is given to suggest the heath care measures for reducing the transmissibility of COVID-19 infection in the population.

## Data Availability

All data is available online

